# Associations between cardiorespiratory fitness and lifestyle-related factors with DNA methylation-based aging clocks in older men: WASEDA’S Health Study

**DOI:** 10.1101/2023.04.12.23288187

**Authors:** Takuji Kawamura, Zsolt Radak, Hiroki Tabata, Hiroshi Akiyama, Nobuhiro Nakamura, Ryoko Kawakami, Tomoko Ito, Chiyoko Usui, Matyas Jokai, Ferenc Torma, Hyeon-Ki Kim, Motohiko Miyachi, Suguru Torii, Katsuhiko Suzuki, Kaori Ishii, Shizuo Sakamoto, Koichiro Oka, Mitsuru Higuchi, Isao Muraoka, Kristen M. McGreevy, Steve Horvath, Kumpei Tanisawa

## Abstract

DNA methylation-based age estimators (DNAm aging clocks) are currently one of the most promising biomarkers for predicting biological age. However, the relationships between objectively measured physical fitness, including cardiorespiratory fitness, and DNAm aging clocks are largely unknown. We investigated the relationships between physical fitness and the age-adjusted value from the residuals of the regression of DNAm aging clock to chronological age (DNAmAgeAcceleration: DNAmAgeAccel) and attempted to determine the relative contribution of physical fitness variables to DNAmAgeAccel in the presence of other lifestyle factors. DNA samples from 144 Japanese men aged 65–72 years were used to calculate first- (*i.e.*, DNAmHorvath and DNAmHannum) and second- (*i.e.*, DNAmPhenoAge, DNAmGrimAge and DNAmFitAge) generation DNAm aging clocks. Various surveys and measurements were conducted, including physical fitness, body composition, blood biochemical parameters, nutrients intake, smoking, alcohol consumption, disease status, sleep status, and chronotype. The peak oxygen uptake (VO_2peak_) per kg body weight had a significant negative correlation with GrimAgeAccel (*r* = -0.222, *p* = 0.008). A comparison of the tertile groups showed that the GrimAgeAccel of the highest VO_2peak_ group was decelerated by 1.6 years compared to the lowest group (*p* = 0.035). Multiple regression analysis suggested that rather than physical fitness, serum triglycerides, carbohydrate intake, and smoking status, were significantly associated with DNAmAgeAccel. In conclusion, the contribution of cardiorespiratory fitness to DNAmAgeAccel was relatively low compared to lifestyle factors such as smoking. However, this study reveals a negative relationship between cardiorespiratory fitness and DNAmAgeAccel in older men.

## 1. Introduction

Population aging is an issue in many countries around the world; in fact, between 2000 and 2019, the global life expectancy at birth increased by 6.5 years (World Health Organization, 2022). More importantly, there is a considerable gap of approximately 10 years between life expectancy and healthy life expectancy, regardless of the region and wealth of countries (World Health Organization, 2022). This gap is not limited to individual problems, such as a decline in quality of life but is also directly linked to social and economic losses, such as increased medical costs and long-term care burdens. To improve this current situation and to extend health span, it is required to establish intervention strategies that target aging itself, the strongest risk factor for each disease, rather than just focusing on organ- and disease-based segmented medicine (Campisi et al., 2019). Chronological age, the number of years since birth, is frequently used as a measure of aging but is often accompanied by large inter-individual variability on health outcomes in older adults based on this measure. Therefore, aging biomarkers that can predict biological age, which reflect the biological state of the individual, have been explored for over 30 years (Baker & Sprott, 1988). Developing such aging surrogate biomarkers could be useful tools for assessing and validating anti-aging interventions on the rate of aging in each individual within a relatively short time frame.

DNA methylation-based age estimators (DNAm aging clocks) are currently considered one of the most promising biomarkers for predicting biological age (Jylhävä et al., 2017). Briefly, the majority of DNAm aging clocks are calculated by (1) selecting key Cytosine-Phosphate-Guanine (CpG) sites where hyper- and hypo-methylation correlate with age and other phenotypes and weighting them with a linear model and (2) creating an equation to estimate age based on the methylation level of each CpG site (Simpson & Chandra, 2021). For example, the ‘first generation’ DNAm aging clocks, the DNAmAgeHorvath and the DNAmAgeHannum, are calculated by methylation levels of 353 and 71 CpG sites, respectively, and have been shown to correlate strongly with chronological age (Hannum et al., 2013; Horvath, 2013). The subsequently proposed ‘second generation’ composite clocks, DNAmPhenoAge and DNAmGrimAge, use DNAm to predict non-DNA traits, which can be used as additional variables to predict biological age, disease status, and mortality accurately (Levine et al., 2018; Lu et al., 2019; Simpson & Chandra, 2021). Recently, a novel composite biomarker, DNAmFitAge, was developed that incorporates the physical fitness parameters of gait speed, hand grip strength, forced expiratory volume in one second (FEV1), and maximal oxygen uptake (VO_2max_) (McGreevy et al., 2023). These DNAm-based biomarkers are expressed as absolute values (DNAmAge) or age-adjusted values from the residuals of the regression of each DNAm aging clock to chronological age (DNAmAgeAcceleration: DNAmAgeAccel), and the main advantage of these biomarkers is to be able to quantify the individual biological aging. Therefore, targeting these DNAm aging clocks and identifying which lifestyle habits slow down or reverse the aging processes would contribute to promoting healthy aging.

At present, several potential ‘geroprotector’ candidates (including rapamycin, sirtuin-activating compounds, and metformin) are emerging in the field of aging biology, and several clinical trials are being initiated based on the findings of model organisms (Kulkarni et al., 2022; Partridge et al., 2020); however, the validation of these pharmacological approaches is still insufficient. In contrast, exercise is a well-documented effective ‘geroprotector’ that reduces functional disability and age-related diseases and extends health span (Chakravarty et al., 2008; Lear et al., 2017; Moore et al., 2012). The relationships between respective DNAmAgeAccel and physical activity and functions have been investigated, although there is limited evidence to support this. Specifically, physical activity level seems to negatively correlate with second-generation clocks (*i.e.*, DNAmPhenoAge and DNAmGrimAge) rather than with first-generation clocks (*i.e.*, DNAmAgeHorvath and DNAmAgeHannum) (Kankaanpää et al., 2021; Kim et al., 2022; Kresovich et al., 2021; Levine et al., 2018; Lu et al., 2019; McCrory et al., 2021; Quach et al., 2018; Sillanpää et al., 2019; Stevenson et al., 2019; Zhao et al., 2019). However, whether DNAmAgeAccel is related to physical functions such as grip strength, walking speed, and pulmonary function is controversial (Belsky et al., 2018; Föhr et al., 2022; Maddock et al., 2020; Marioni et al., 2015; McCrory et al., 2021; Petersen et al., 2021; Sillanpaä et al., 2018; Sillanpää et al., 2021; Simpkin et al., 2017). In addition, there is insufficient evidence on the relationships between DNAmAgeAccel and VO_2max_, a strong predictor of all-cause mortality (Jokai et al., 2022; Sillanpää et al., 2021). In sum, it is essential to clarify the relationship between objectively measured physical fitness and DNAmAgeAccel, including VO_2max_ measurements, to develop a basis for establishing an efficient exercise intervention against biological aging. Regarding healthy lifestyle habits to slow down DNAmAgeAccel, lifestyle-related variables other than physical fitness should be included naturally. In fact, lifestyle-related variables such as body composition (Horvath et al., 2014; Levine et al., 2018; Lu et al., 2019; McCartney et al., 2018; Quach et al., 2018), diet (Levine et al., 2018; Lu et al., 2019; Quach et al., 2018), smoking (Kim et al., 2022; Levine et al., 2018; Lu et al., 2019), alcohol consumption (Kim et al., 2022; Quach et al., 2018), sleep status (Carroll et al., 2017; Carskadon et al., 2019; Freni-Sterrantino et al., 2022), and blood biochemical parameters (Levine et al., 2018; Lu et al., 2019; Quach et al., 2018) are associated with acceleration or deceleration of DNAm aging clocks.

Therefore, this study primarily aims to determine the relationships between objectively measured physical fitness, including peak oxygen uptake (VO_2peak_), and DNAmAgeAccel in older men. The secondary aim of this study is to determine the relative contribution of lifestyle related variables other than physical fitness to DNAmAgeAccel. To achieve this purpose, we calculated five DNAm aging clocks: DNAmAgeHorvath, DNAmAgeHannum, DNAmPhenoAge, DNAmGrimAge, and DNAmFitAge, and their AgeAccel using DNA samples from 144 Japanese men aged 65–72 years who participated in the Waseda Alumni’s Sports, Exercise, Daily Activity, Sedentariness, and Health Study (WASEDA’S Health Study). Along with these five DNAm aging clocks, we conducted various surveys and measurements, including physical fitness, body composition, blood biochemical parameters, nutrients intake, smoking, alcohol consumption, disease status, sleep status, and chronotype. Through these surveys and measurements, we aim to identify lifestyle-related variables, including physical fitness, associated with biological aging and provide evidence for selecting targets for anti-aging intervention strategies.

## 2. Results

### 2.1. Characteristics of the participants

The characteristics of all participants are shown in **Table 1 and S1**. The overall number of participants in this study was 144 men. The mean chronological age was 68.0 ± 1.9 years, and all participants were Japanese.

**Table 1.** Characteristics of the participants and tertiles of each continuous variable.

| | Mean ± SD | | | *n* |
| --- | --- | --- | --- | --- |
| ***Anthropometric variables*** |  |  |  |  |
| Height (cm) | 168.0 | ± | 5.8 | 144 |
| Body weight (kg) | 66.1 | ± | 8.2 | 144 |
| BMI (kg/m<sup>2</sup>) | 23.4 | ± | 2.5 | 144 |
| Body fat (%) | 21.2 | ± | 5.4 | 144 |
| Fat free mass (kg) | 51.8 | ± | 4.6 | 144 |
| Visceral fat area (cm<sup>2</sup>) | 96.5 | ± | 45.6 | 139 |
| Subcutaneous fat area (cm<sup>2</sup>) | 123.6 | ± | 48.3 | 139 |
| Abdominal circumference (cm) | 84.8 | ± | 7.6 | 144 |
| Calf circumference (cm) | 36.6 | ± | 2.1 | 136 |
| ***Physical fitness variables and blood pressure*** |  |  |  |  |
| VO<sub>2</sub> at VT (mL/min) | 1001.3 | ± | 208.5 | 142 |
| VO<sub>2</sub>/kg at VT (mL/kg/min) | 15.3 | ± | 3.4 | 142 |
| VO<sub>2</sub> at peak (mL/min) | 1736.5 | ± | 293.1 | 144 |
| VO<sub>2</sub>/kg at peak (mL/kg/min) | 26.5 | ± | 4.6 | 144 |
| Grip strength (kg) | 35.2 | ± | 5.7 | 143 |
| Leg extension power (W) | 1062.4 | ± | 265.1 | 143 |
| SBP (mmHg) | 138.8 | ± | 20.4 | 144 |
| DBP (mmHg) | 84.3 | ± | 12.1 | 144 |
| ***Blood biochemical variables*** |  |  |  |  |
| Insulin (μU/mL) | 6.5 | ± | 4.2 | 144 |
| Glucose (mg/dL) | 104.5 | ± | 18.1 | 144 |
| HbA1c (%) | 5.7 | ± | 0.5 | 144 |
| TG (mg/dL) | 111.7 | ± | 62.6 | 144 |
| Total-C (mg/dL) | 214.3 | ± | 31.9 | 144 |
| HDL-C (mg/dL) | 63.6 | ± | 16.1 | 144 |
| LDL-C (mg/dL) | 124.5 | ± | 29.2 | 144 |
| ***Nutrients variables*** |  |  |  |  |
| Energy intake (kcal/day) | 2106 | ± | 558 | 144 |
| Protein (g/1000 kcal/day) | 42.0 | ± | 7.4 | 144 |
| Fat (g/1000 kcal/day) | 32.0 | ± | 6.9 | 144 |
| CHO (g/1000 kcal/day) | 117.8 | ± | 21.9 | 144 |
| Fe (mg/1000 kcal/day) | 4.9 | ± | 1.2 | 144 |
| Zn (mg/1000 kcal/day) | 4.6 | ± | 0.7 | 144 |
| Cu (mg/1000 kcal/day) | 0.7 | ± | 0.14 | 144 |
| Mn (mg/1000 kcal/day) | 1.6 | ± | 0.4 | 144 |
| Vitamin A (μg RAE/1000 kcal/day) | 516 | ± | 268 | 144 |
| Vitamin C (mg/1000 kcal/day) | 74.2 | ± | 31.2 | 144 |
| α-tocopherol (mg/1000 kcal/day) | 4.6 | ± | 1.1 | 144 |
| β-carotene (μg/1000 kcal/day) | 2335 | ± | 1385 | 144 |
| ***DNAmAge and DNAmAgeAccel*** |  |  |  |  |
| Chronological Age (years) | 68.0 | ± | 1.9 | 144 |
| DNAmAgeHorvath (predicted years) | 62.2 | ± | 4.5 | 144 |
| DNAmAgeHannum (predicted years) | 55.1 | ± | 3.8 | 144 |
| DNAmPhenoAge (predicted years) | 56.7 | ± | 5.5 | 144 |
| DNAmGrimAge (predicted years) | 69.2 | ± | 3.4 | 144 |
| DNAmFitAge (predicted years) | 71.7 | ± | 3.5 | 144 |
| HorvathAgeAccel (residuals) | 0 | ± | 4.4 | 144 |
| HannumAgeAccel (residuals) | 0 | ± | 3.7 | 144 |
| PhenoAgeAccel (residuals) | 0 | ± | 5.5 | 144 |
| GrimAgeAccel (residuals) | 0 | ± | 3.2 | 144 |
| FitAgeAccel (residuals) | 0 | ± | 3.3 | 144 |

Data are mean ± standard deviation (SD). BMI, body mass index; VT, ventilatory threshold; SBP, systolic blood pressure; DBP, diastolic blood pressure; HbA1c, Hemoglobin A1c; TG, triglyceride; Total-C, total cholesterol; HDL-C, high-density lipoprotein cholesterol; LDL-C, low-density lipoprotein cholesterol; CHO, carbohydrate; Fe, iron; Zn, zinc; Cu, copper; Mn, manganese; Accel, acceleration.

### 2.2. Chronological age, DNA methylation age, and DNA methylation age acceleration

The correlations between chronological age and DNAmAge is shown in **Figure 1a**. Chronological age significantly correlated with DNAmAgeHannum (*r* = 0.230, *p* < 0.01), DNAmGrimAge (*r* = 0.380, *p* < 0.001), and DNAmFitAge (*r* = 0.370, *p* < 0.001), respectively, while there was no significant correlation with DNAmAgeHorvath (*r* = 0.083, *p* = 0.321) and DNAmPhenoAge (*r* = 0.057, *p* = 0.501).

**Figure 1.**
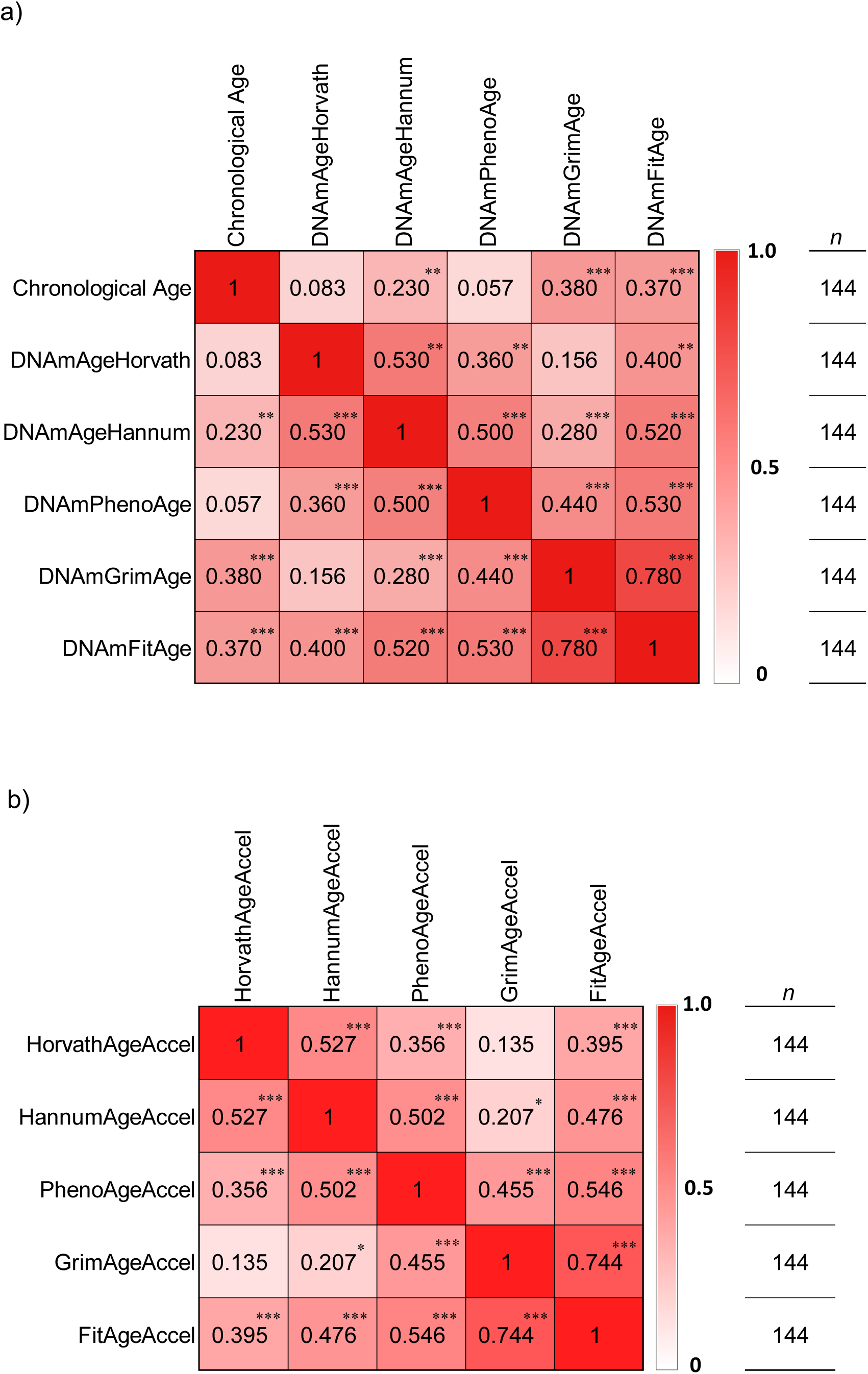
Correlations between chronological age, DNA methylation aging clock, and DNA methylation age acceleration. a) Correlations between chronological age and each DNA methylation aging clock. b) Correlations between each DNA methylation age acceleration. Significant correlations at *p* < 0.05, *p* < 0.01 and *p* < 0.001 are indicated by *, ** and ***, respectively.

The correlations between each DNAmAgeAccel is shown in **Figure 1b**. Except for HorvathAgeAccel and GrimAgeAccel, rest of the DNAmAgeAccel significantly correlated with each other (HorvathAgeAccel × HannumAgeAccel: *r* = 0.527, *p* < 0.001; HorvathAgeAccel × PhenoAgeAccel: *r* = 0.356, *p* < 0.001; HorvathAgeAccel × FitAgeAccel: *r* = 0.395, *p* < 0.001; HannumAgeAccel ×PhenoAgeAccel: *r* = 0.502, *p* < 0.001; HannumAgeAccel × GrimAgeAccel: *r* = 0.207, *p* = 0.013; HannumAgeAccel ×FitAgeAccel: *r* = 0.476, *p* < 0.001; PhenoAgeAccel × GrimAgeAccel: *r* = 0.455, *p* < 0.001; PhenoAgeAccel × FitAgeAccel: *r* = 0.546, *p* < 0.001; GrimAgeAccel × FitAgeAccel: *r* = 0.744, *p* < 0.001).

In this study, relatively high correlation coefficients were observed between chronological age and second generation DNAmAge (*i.e.*, DNAmGrimAge and DNAmFitAge). Similarly, relatively high correlation coefficients were seen among the second generation DNAmAgeAccel. Based on the results of these correlation analyses, we present the data on the relationships between PhenoAgeAccel, GrimAgeAccel, FitAgeAccel, and other lifestyle-related variables.

### 2.3. DNA methylation age acceleration and physical fitness variables

The correlations between DNAmAgeAccel and physical fitness is shown in **Figure 2a**. VO_2_/kg at ventilatory threshold (VT) and VO_2_/kg at Peak significantly correlated with GrimAgeAccel (VO_2_/kg at VT: *r* = -0.232, *p* = 0.005; VO_2_/kg at Peak: *r* = -0.222, *p* = 0.008) (**Figure 2b and 2c**). Chronological age significantly correlated with VO_2_ at VT (*r* = -0.184, *p* = 0.029) and VO_2_ at Peak (*r* = -0.175, *p* = 0.036), respectively (**Figure 2a)**.

**Figure 2.**
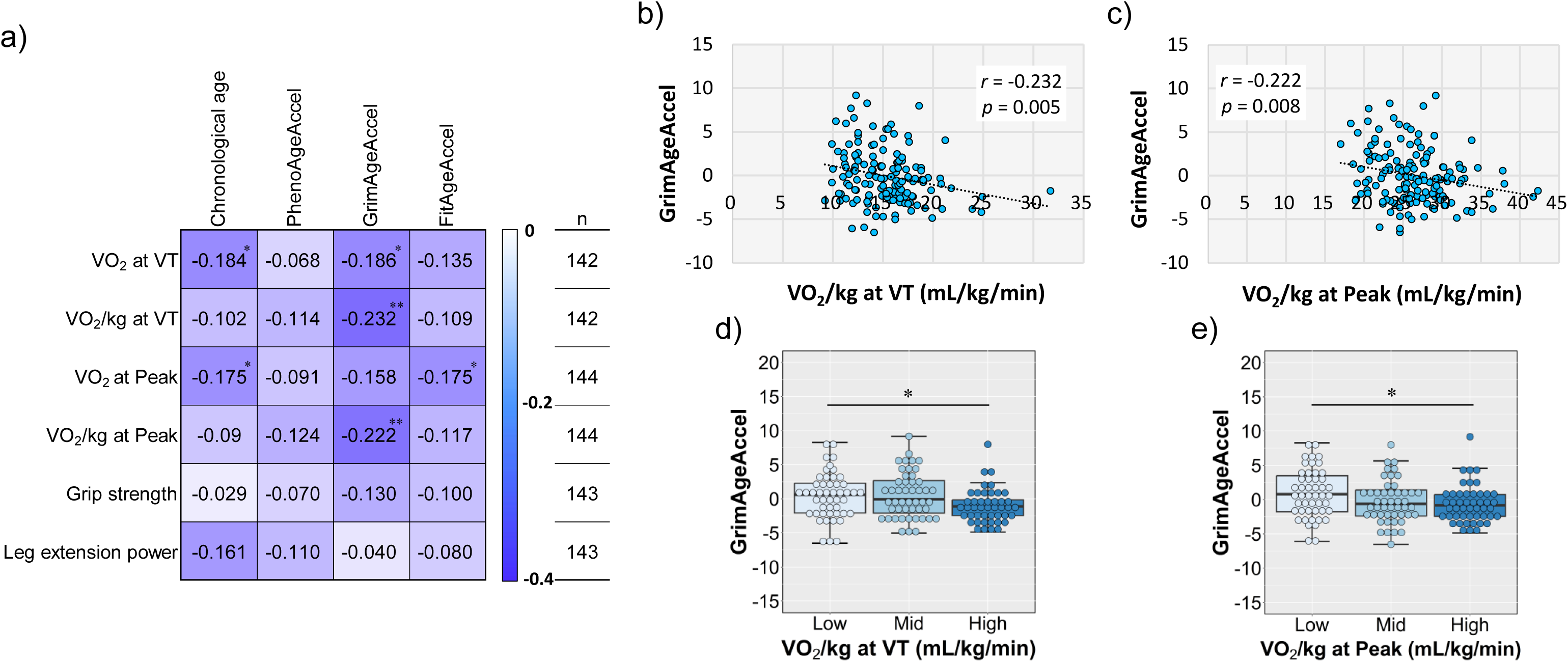
Associations between physical fitness variables and DNA methylation age acceleration. a) Correlations between physical fitness variables and DNA methylation age acceleration. b, c) Scatter plots of GrimAgeAccel and VO_2_ per body weight at ventilatory threshold and Peak. d, e) Comparison of GrimAgeAccel in tertile groups of VO_2_ per body weight at ventilatory threshold and Peak. VO_2_, oxygen uptake; VT, ventilatory threshold. Significant correlations or differences at *p* < 0.05 and *p* < 0.01 are indicated by * and **, respectively.

### 2.4. DNA methylation age acceleration and anthropometric variables

The correlations between DNAmAgeAccel and anthropometric variables are shown in **Figure 3a**. Body fat significantly correlated with PhenoAgeAccel (*r* = 0.205, *p* = 0.014, **Figure 3b**) and GrimAgeAccel (*r* = 0.188 *p* = 0.024, **Figure 3c**), respectively. Visceral fat area significantly correlated with PhenoAgeAccel (*r* = 0.215, *p* = 0.011, **Figure 3d**) and GrimAgeAccel (*r* = 0.251, *p* = 0.003, **Figure 3e**), respectively. Fat free mass and calf circumference significantly correlated with FitAgeAccel (fat free mass: *r* = -0.167, *p* = 0.045; calf circumference: *r* = -0.216, *p* = 0.012, **Figure 3f** and **3g**), respectively. Chronological age significantly correlated with subcutaneous fat area (*r* = -0.194, *p* = 0.022) (**Figure 3a**).

**Figure 3.**
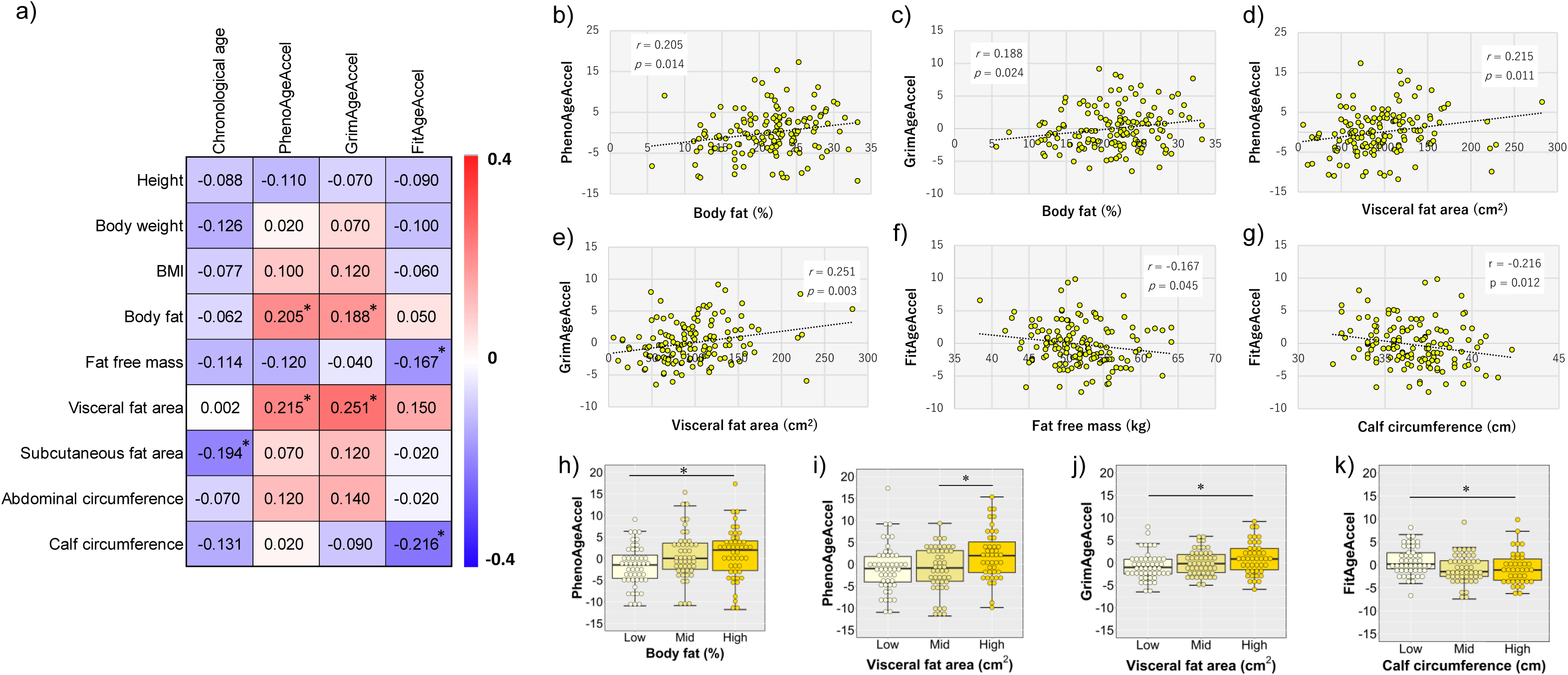
Associations between anthropometric variables and DNA methylation age acceleration. a) Correlations between anthropometric variables and DNA methylation age acceleration. b-g) Scatter plots of each DNA methylation age acceleration and body fat, visceral fat area, fat free mass, and calf circumference. h-k) Comparison of each DNA methylation age acceleration in tertile groups of body fat, visceral fat area, and calf circumference. BMI, body mass index. Significant correlations or difference at *p* < 0.05 is indicated by *.

### 2.5. DNA methylation age acceleration and biochemical variables

The correlations between DNAmAgeAccel and blood biochemical variables are shown in **Figure 4a**. PhenoAgeAccel significantly correlated with triglycerides (TG) (*r* = 0.178, *p* = 0.032, **Figure 4b**), high-density lipoprotein cholesterol (HDL-C) (*r* = -0.221, *p* = 0.008, **Figure 4e**), respectively. GrimAgeAccel significantly correlated with TG (*r* = 0.277, *p* < 0.001, **Figure 4c**), HDL-C (*r* = -0.183, *p* = 0.028, **Figure 4f**), respectively. FitAgeAccel significantly correlated with TG (*r* = 0.267, *p* = 0.001, **Figure 4d**). However, there was no significant correlation between chronological age and any of the variables (**Figure 4a**).

**Figure 4.**
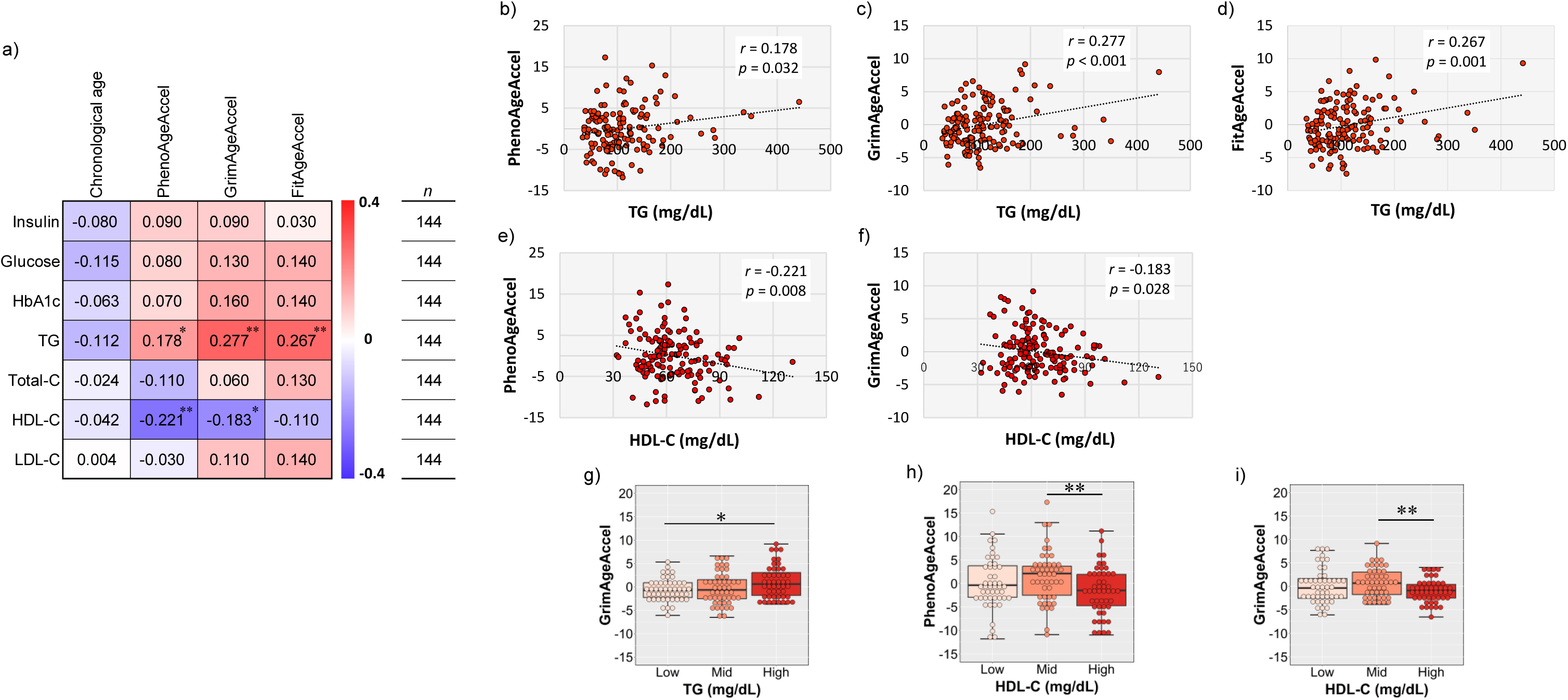
Associations between blood biochemical variables and DNA methylation age acceleration. a) Correlations between blood biochemical variables and DNA methylation age acceleration. b-f) Scatter plots of each DNA methylation age acceleration and triglyceride (TG), high-density lipoprotein cholesterol (HDL-C). g-i) Comparison of each DNA methylation age acceleration in tertile groups of TG, HDL-C. HbA1c, Hemoglobin A1c; Total-C, total cholesterol; LDL-C, low-density lipoprotein cholesterol. Significant correlations or differences at *p* < 0.05 and *p* < 0.01 are indicated by * and **, respectively.

### 2.6. DNA methylation age acceleration and nutrients variables

The correlations between DNAmAgeAccel and nutrients variables are shown in **Figure 5a**. PhenoAgeAccel significantly correlated with Cu (*r* = -0.166, *p* = 0.047, **Figure 5e**), vitamin C (*r* = -0.204, *p* = 0.014, **Figure 5h**), and *β*-carotene (*r* = -0.181, *p* = 0.030, **Figure 5j**), respectively. GrimAgeAccel significantly correlated with CHO (*r* = -0.260, *p* = 0.002, **Figure 5b**), Fe (*r* = -0.168, *p* = 0.044, **Figure 5c**), Cu (*r* = -0.284, *p* < 0.001, **Figure 5f**), vitamin C (*r* = -0.225, *p* = 0.007, **Figure 5i**), and *β*-carotene (*r* = -0.222, *p* = 0.008, **Figure 5k**), respectively. FitAgeAccel significantly correlated with Fe (*r* = - 0.187, *p* = 0.025, **Figure 5d**), Cu (*r* = -0.264, *p* < 0.001, **Figure 5g**), and *β*-carotene (*r* = -0.172, *p* = 0.039, **Figure 5l**), respectively. Chronological age significantly correlated with vitamin C (*r* = 0.246, *p* = 0.003) and *β*-carotene (*r* = 0.197, *p* = 0.018), respectively (**Figure 5a)**.

**Figure 5.**
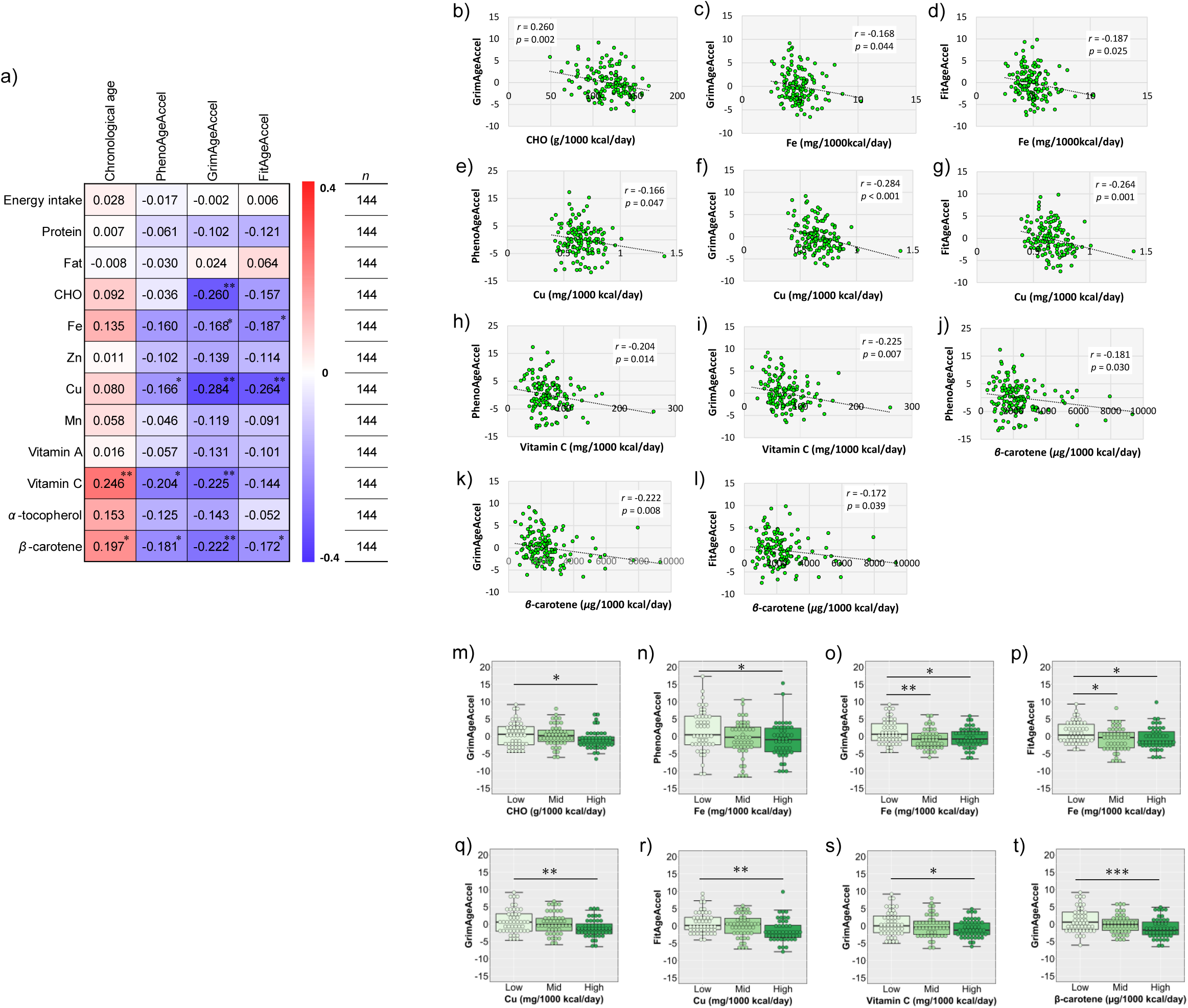
Associations between nutrients variables and DNA methylation age acceleration. a) Correlations between nutrients variables and DNA methylation age acceleration. b-l) Scatter plots of each DNA methylation age acceleration and carbohydrate (CHO), iron (Fe), copper (Cu), vitamin C, and *β*-carotene. m-t) Comparison of each DNA methylation age acceleration in tertile groups of CHO, Fe, Cu, vitamin C, *β*-carotene. Zn, zinc; Mn, manganese. Significant correlations or differences at *p* < 0.05, *p* < 0.01 and *p* < 0.001 are indicated by *, ** and ***, respectively.

### 2.7. DNA methylation age acceleration of the participants by tertile of each continuous variable

The characteristics of participants by tertile for each continuous variable are shown in **Table S2**. The DNAmAgeAccel for each of these tertile groups can also be found in **Tables S3**–**S5**. The only variables for which statistically significant differences were found are described below. Among the physical fitness variables, PhenoAgeAccel was slower at T3 than at T2 for leg extension (*p* = 0.017). GrimAgeAccel were slower at T3 than at T1 for VO_2_/kg at VT (*p* = 0.042, **Figure 2d**) and VO_2_/kg at Peak (*p* = 0.035, **Figure 2e**). Among the anthropometric variables, PhenoAgeAccel was faster at T3 than at T1 for body fat composition (*p* = 0.029, **Figure 3h**) and was faster at T3 than at T2 for visceral fat area (*p* = 0.036, **Figure 3i**). GrimAgeAccel was faster at T3 than at T1 for visceral fat area (*p* = 0.018, **Figure 3j**). FitAgeAccel was slower at T3 than at T1 for calf circumference (*p* = 0.023, **Figure 3k**). Among the blood biochemical variables, GrimAgeAccel were faster at T3 than at T1 for TG (*p* = 0.030, **Figure 4g**). PhenoAgeAccel was slower at T3 than at T2 for HDL-C (*p* = 0.010, **Figure 4h**). GrimAgeAccel was slower at T3 than T2 for HDL-C *(p* = 0.006, **Figure 4i**). Among the nutrients variables, PhenoAgeAccel was slower at T3 than at T1 for Fe (*p* = 0.045, **Figure 5n**). GrimAgeAccel was slower at T3 than at T1 for CHO (*p* = 0.048, **Figure 5m**), Cu (*p* = 0.004, **Figure 5q**), vitamin C (*p* = 0.039, **Figure 5s**), and *β*-carotene (*p* < 0.001, **Figure 5t**). GrimAgeAccel was slower at T2 (*p* = 0.006) and T3 (*p* = 0.020, **Figure 5o**) than at T1 for Fe. FitAgeAccel was slower at T2 (*p* = 0.019) and T3 (*p* = 0.034, **Figure 5p**) than at T1 for Fe and was slower at T3 than at T1 for Cu (*p* = 0.001, **Figure 5r**).r

**Table 2.**
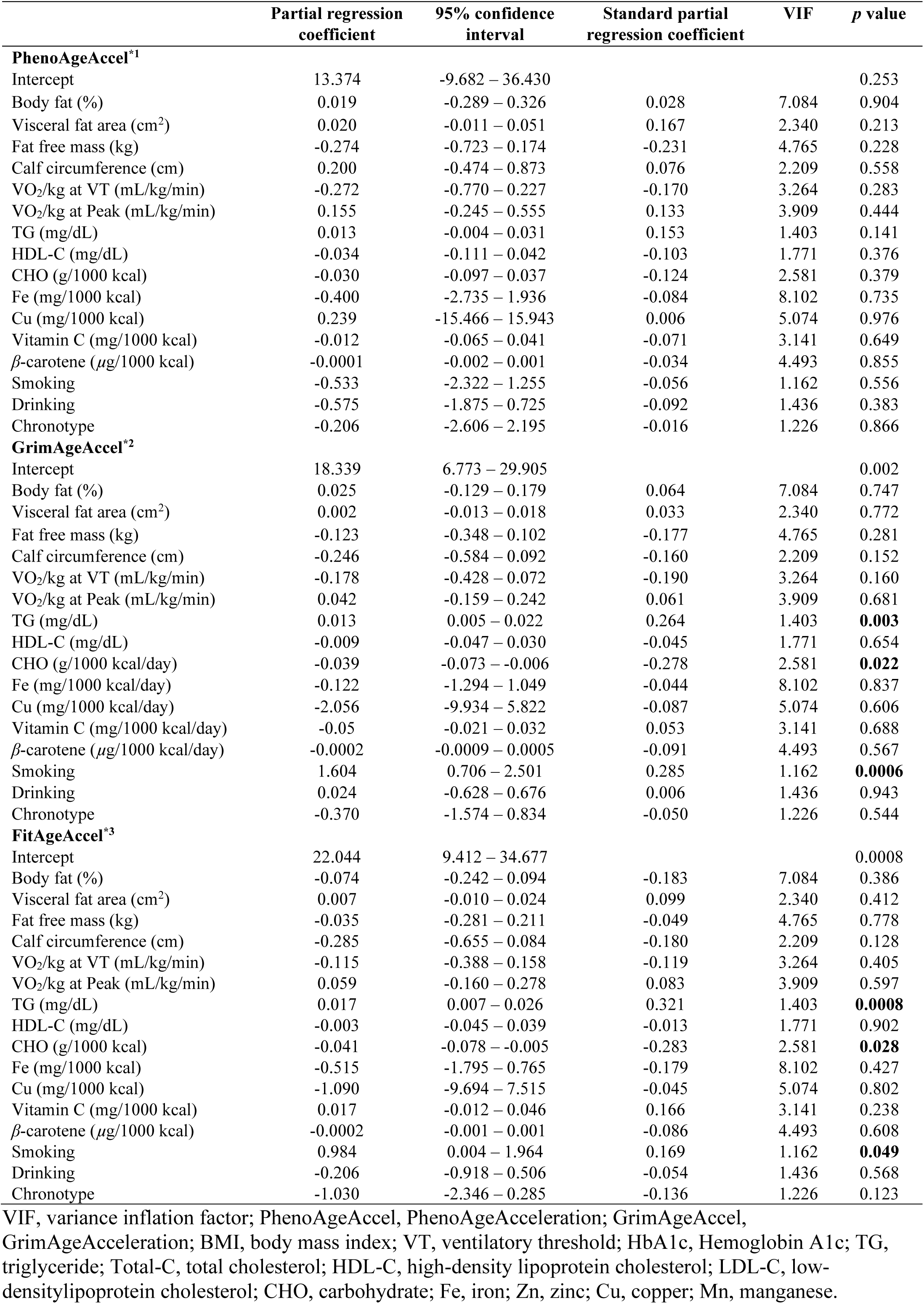
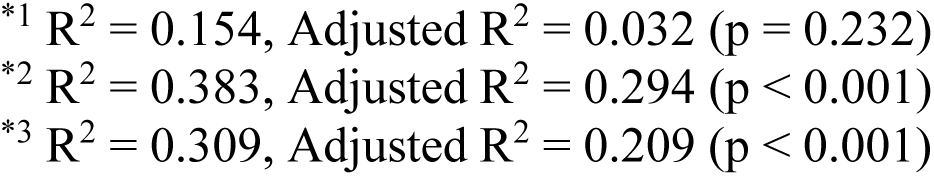
Multiple regression analysis of each DNA methylation age acceleration.

|  | Partial regression<br>coefficient | 95% confidence<br>interval | Standard partial<br>regression coefficient | VIF | *p* value |
| --- | --- | --- | --- | --- | --- |
| **PhenoAgeAccel<sup>\*1</sup>** |  |  |  |  |  |
| Intercept | 13.374 | -9.682 – 36.430 |  |  | 0.253 |
| Body fat (%) | 0.019 | -0.289 – 0.326 | 0.028 | 7.084 | 0.904 |
| Visceral fat area (cm<sup>2</sup>) | 0.020 | -0.011 – 0.051 | 0.167 | 2.340 | 0.213 |
| Fat free mass (kg) | -0.274 | -0.723 – 0.174 | -0.231 | 4.765 | 0.228 |
| Calf circumference (cm) | 0.200 | -0.474 – 0.873 | 0.076 | 2.209 | 0.558 |
| VO<sub>2</sub>/kg at VT (mL/kg/min) | -0.272 | -0.770 – 0.227 | -0.170 | 3.264 | 0.283 |
| VO<sub>2</sub>/kg at Peak (mL/kg/min) | 0.155 | -0.245 – 0.555 | 0.133 | 3.909 | 0.444 |
| TG (mg/dL) | 0.013 | -0.004 – 0.031 | 0.153 | 1.403 | 0.141 |
| HDL-C (mg/dL) | -0.034 | -0.111 – 0.042 | -0.103 | 1.771 | 0.376 |
| CHO (g/1000 kcal) | -0.030 | -0.097 – 0.037 | -0.124 | 2.581 | 0.379 |
| Fe (mg/1000 kcal) | -0.400 | -2.735 – 1.936 | -0.084 | 8.102 | 0.735 |
| Cu (mg/1000 kcal) | 0.239 | -15.466 – 15.943 | 0.006 | 5.074 | 0.976 |
| Vitamin C (mg/1000 kcal) | -0.012 | -0.065 – 0.041 | -0.071 | 3.141 | 0.649 |
| β-carotene (μg/1000 kcal) | -0.0001 | -0.002 – 0.001 | -0.034 | 4.493 | 0.855 |
| Smoking | -0.533 | -2.322 – 1.255 | -0.056 | 1.162 | 0.556 |
| Drinking | -0.575 | -1.875 – 0.725 | -0.092 | 1.436 | 0.383 |
| Chronotype | -0.206 | -2.606 – 2.195 | -0.016 | 1.226 | 0.866 |
| **GrimAgeAccel<sup>\*2</sup>** |  |  |  |  |  |
| Intercept | 18.339 | 6.773 – 29.905 |  |  | 0.002 |
| Body fat (%) | 0.025 | -0.129 – 0.179 | 0.064 | 7.084 | 0.747 |
| Visceral fat area (cm<sup>2</sup>) | 0.002 | -0.013 – 0.018 | 0.033 | 2.340 | 0.772 |
| Fat free mass (kg) | -0.123 | -0.348 – 0.102 | -0.177 | 4.765 | 0.281 |
| Calf circumference (cm) | -0.246 | -0.584 – 0.092 | -0.160 | 2.209 | 0.152 |
| VO<sub>2</sub>/kg at VT (mL/kg/min) | -0.178 | -0.428 – 0.072 | -0.190 | 3.264 | 0.160 |
| VO<sub>2</sub>/kg at Peak (mL/kg/min) | 0.042 | -0.159 – 0.242 | 0.061 | 3.909 | 0.681 |
| TG (mg/dL) | 0.013 | 0.005 – 0.022 | 0.264 | 1.403 | **0.003** |
| HDL-C (mg/dL) | -0.009 | -0.047 – 0.030 | -0.045 | 1.771 | 0.654 |
| CHO (g/1000 kcal/day) | -0.039 | -0.073 – -0.006 | -0.278 | 2.581 | **0.022** |
| Fe (mg/1000 kcal/day) | -0.122 | -1.294 – 1.049 | -0.044 | 8.102 | 0.837 |
| Cu (mg/1000 kcal/day) | -2.056 | -9.934 – 5.822 | -0.087 | 5.074 | 0.606 |
| Vitamin C (mg/1000 kcal/day) | -0.05 | -0.021 – 0.032 | 0.053 | 3.141 | 0.688 |
| β-carotene (μg/1000 kcal/day) | -0.0002 | -0.0009 – 0.0005 | -0.091 | 4.493 | 0.567 |
| Smoking | 1.604 | 0.706 – 2.501 | 0.285 | 1.162 | **0.0006** |
| Drinking | 0.024 | -0.628 – 0.676 | 0.006 | 1.436 | 0.943 |
| Chronotype | -0.370 | -1.574 – 0.834 | -0.050 | 1.226 | 0.544 |
| **FitAgeAccel<sup>\*3</sup>** |  |  |  |  |  |
| Intercept | 22.044 | 9.412 – 34.677 |  |  | 0.0008 |
| Body fat (%) | -0.074 | -0.242 – 0.094 | -0.183 | 7.084 | 0.386 |
| Visceral fat area (cm<sup>2</sup>) | 0.007 | -0.010 – 0.024 | 0.099 | 2.340 | 0.412 |
| Fat free mass (kg) | -0.035 | -0.281 – 0.211 | -0.049 | 4.765 | 0.778 |
| Calf circumference (cm) | -0.285 | -0.655 – 0.084 | -0.180 | 2.209 | 0.128 |
| VO<sub>2</sub>/kg at VT (mL/kg/min) | -0.115 | -0.388 – 0.158 | -0.119 | 3.264 | 0.405 |
| VO<sub>2</sub>/kg at Peak (mL/kg/min) | 0.059 | -0.160 – 0.278 | 0.083 | 3.909 | 0.597 |
| TG (mg/dL) | 0.017 | 0.007 – 0.026 | 0.321 | 1.403 | **0.0008** |
| HDL-C (mg/dL) | -0.003 | -0.045 – 0.039 | -0.013 | 1.771 | 0.902 |
| CHO (g/1000 kcal) | -0.041 | -0.078 – -0.005 | -0.283 | 2.581 | **0.028** |
| Fe (mg/1000 kcal) | -0.515 | -1.795 – 0.765 | -0.179 | 8.102 | 0.427 |
| Cu (mg/1000 kcal) | -1.090 | -9.694 – 7.515 | -0.045 | 5.074 | 0.802 |
| Vitamin C (mg/1000 kcal) | 0.017 | -0.012 – 0.046 | 0.166 | 3.141 | 0.238 |
| β-carotene (μg/1000 kcal) | -0.0002 | -0.001 – 0.001 | -0.086 | 4.493 | 0.608 |
| Smoking | 0.984 | 0.004 – 1.964 | 0.169 | 1.162 | **0.049** |
| Drinking | -0.206 | -0.918 – 0.506 | -0.054 | 1.436 | 0.568 |
| Chronotype | -1.030 | -2.346 – 0.285 | -0.136 | 1.226 | 0.123 |

VIF, variance inflation factor; PhenoAgeAccel, PhenoAgeAcceleration; GrimAgeAccel, GrimAgeAcceleration; BMI, body mass index; VT, ventilatory threshold; HbA1c, Hemoglobin A1c; TG, triglyceride; Total-C, total cholesterol; HDL-C, high-density lipoprotein cholesterol; LDL-C, low-density lipoprotein cholesterol; CHO, carbohydrate; Fe, iron; Zn, zinc; Cu, copper; Mn, manganese.
\*<sup>1</sup> $R^2 = 0.154$ , Adjusted $R^2 = 0.032$ ( $p = 0.232$ )
\*<sup>2</sup> $R^2 = 0.383$ , Adjusted $R^2 = 0.294$ ( $p < 0.001$ )
\*<sup>3</sup> $R^2 = 0.309$ , Adjusted $R^2 = 0.209$ ( $p < 0.001$ )

### 2.8. DNA methylation age acceleration of the categorical variables

The characteristics of the participant’s categorical variables are shown in **Table S1**, and the DNAmAgeAccel for each of categorical variables is shown in **Figure 6** and **Tables S6**–**S10,** respectively. Regarding smoking status, current smokers had faster GrimAgeAccel (*p* < 0.001) and FitAgeAccel (*p* = 0.007) than non-smokers (**Figure 6a**, **6b** and **Table S6**). The GrimAgeAccel of past smokers was also faster than that of non-smokers (*p* = 0.030). Furthermore, the GrimAgeAccel of current smokers was faster than that of past smokers (*p* < 0.001). Correlation analysis of the number of cigarettes smoked and the years of smoking with each DNAmAgeAccel showed a significant correlation between the years of smoking and the GrimAgeAccel (*r* = 0.478, *p* < 0.001) and FitAgeAccel (*r* = 0.326, *p* < 0.001), respectively (**Figure 6e, 6f, 6g**). For drinking status, the group that drank 5-7 times a week had a faster GrimAgeAccel than the group that drank once a week or less (*p* = 0.015) (**Figure 6c, Table S7**). For the Pittsburgh Sleep Quality Index (PSQI) score, there was no difference in any of the DNAmAgeAccel by the presence or absence of sleep disorders (**Table S8**). For the Morningness-Eveningness Questionnaire (MEQ) score, the GrimAgeAccel of the moderate and definite morning type group was significantly slower than the intermediate and moderate evening type group (*p* = 0.033) (**Figure 6d, Table S9**). For each disease, the GrimAgeAccel and FitAgeAccel in the group with dyslipidemia were 1.8–1.9 years earlier than those in the group without the disease, while there was no significant difference in any of the DNAmAgeAccel in other diseases (**Table S10**).

**Figure 6.**
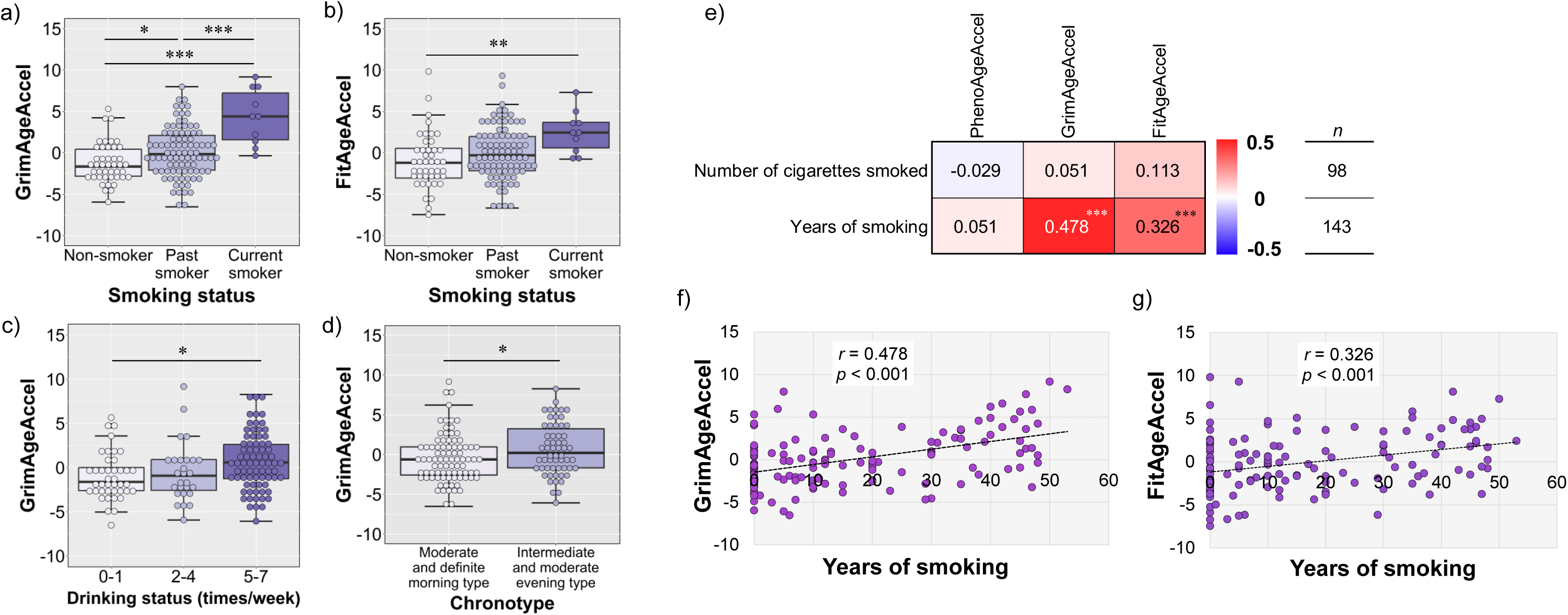
Associations between smoking status, drinking status, chronotype, and DNA methylation age acceleration. a-d) Comparison of each DNA methylation age acceleration in binary and tertile groups of smoking status, drinking status, and chronotype. e) Correlations between number of cigarettes smoked and years of smoking and DNA methylation age acceleration. f, g) Scatter plots of each DNA methylation age acceleration and years of smoking. Significant correlations or differences at *p* < 0.05, *p* < 0.01 and *p* < 0.001 are indicated by *, ** and ***, respectively.

#### Multiple regression analysis of each DNA methylation age acceleration

The results of the multiple regression analysis using the forced entry method for each DNAmAgeAccel are presented in **Table 2**. A significant regression equation was obtained for GrimAgeAccel (*p* < 0.001) and FitAgeAccel (*p* < 0.001), although it was not significant for PhenoAgeAccel (*p* = 0.232). The contribution of the 16 variables entered (adjusted R^2^) was 3.2% for PhenoAgeAccel, 29.4% for GrimAgeAccel, and 20.9% for FitAgeAccel.

## 3. Discussion

The present study revealed that VO_2_/kg at VT and at Peak, fat free mass, calf circumference, serum HDL-C, daily intake of CHO, Fe, Cu, vitamin C, and *β*-carotene were negatively related with DNAmAgeAccel in older men with a narrow age range. In contrast, it also demonstrated that body fat, visceral fat area, and serum TG were positively related to DNAmAgeAccel. Furthermore, frequent alcohol consumption and past- and current-smoking status were associated with accelerated DNAmAgeAccel, while a morning lifestyle was associated with deceleration of it. Multiple regression analysis suggested that rather than physical fitness, serum TG, CHO intake, and smoking status were significantly associated with DNAmAgeAccel.

Aging is a highly complex biological process, and a vast body of knowledge has been accumulated on the hallmarks of aging to date (López-Otín et al., 2013, 2023; Schmauck-Medina et al., 2022). Epigenetic alteration is one of the hallmarks of aging that involves alterations in DNA methylation patterns, post-translational modification of histones, and chromatin remodeling (López-Otín et al., 2013). Of these, the DNAm aging clock, which is based on changes in DNAm patterns, is currently considered to be the most promising biomarker for predicting biological age among the several existing biomarkers (*e.g.*, telomere length, omics-based age estimators) (Horvath & Raj, 2018; Jylhävä et al., 2017). However, evidence on the relationships between these DNAm aging clocks and phenotypic variables is still accumulating, and specifically, the relationships with objectively measured physical fitness, as represented by VO_2peak_, remains unclear.

In this study, negative correlations were observed between VO_2_/kg at VT and VO_2_/kg at Peak and GrimAgeAccel (**Figure 2b** and **2c**). In addition, comparisons across the tertiles indicated that GrimAgeAccel in the high VO_2_/kg at Peak groups (31.5 ± 3.3 mL/kg/min) decelerated by approximately 1.6 years, compared to that of low group (21.8 ± 1.9 mL/kg/min) (**Figure 2e, Table S4**). Similarly, in the recent report, a study of 303 men and women aged 33-88 years, including participants in the World Rowing Masters Regatta in Velence, Hungary, showed that DNAmAgeAccel (*i.e*., FitAgeAccel) was decelerated by 1.5 years in women and 2.0 years in men in the group with higher indirectly calculated VO_2max_ levels (Women: 42.5 ± 7.5 mL/kg/min; Men: 50.2 ± 10.0 mL/kg/min) compared to those with medium-low group (Women: 28.1 ± 5.4 mL/kg/min; Men: 35.1 ± 6.1 mL/kg/min) (Jokai et al., 2022). Although both studies differ in terms of participants, age range, race, and fitness levels, these findings suggest that retaining a high cardiorespiratory fitness is associated with slowing down the biological aging process. From another perspective, our finding may suggest that excessive reductions in cardiorespiratory fitness accelerates DNAmAgeAccel. This is consistent with the concept in exercise science that states that even minimum amount of physical activity can provide health-promoting effects (Arem et al., 2015; Wen et al., 2011). In this study, VO_2_ at Peak as well as at VT showed a negative relationship with DNAmAgeAccel (*i.e.*, GrimAgeAccel) (**Figure 2b-2e**). These results may be useful for safer prediction of older adults’ biological aging without having to continue the measurement up to a maximal load.

Among the anthropometric variables, there were significant positive correlations between body fat, visceral fat area in both PhenoAgeAccel and GrimAgeAccel, respectively (**Figure 3a-e**). In contrast, fat free mass and calf circumference also showed significant negative correlations with FitAgeAccel (**Figure 3a**, **3f** and **3g**). In comparisons among the tertiles, visceral fat area was most related with DNAmAgeAccel (*i.e.*, PhenoAgeAccel and GrimAgeAccel), which was approximately 1.8–2.8 years faster in the highest group than in the low and middle group, respectively (**Figure 3i**, **3j**, **Table S3** and **S4**). Most previous studies investigating the relationships between anthropometric variables and the DNAm aging clocks have used body mass index (BMI) as an indicator, and some systematic reviews have reported a positive relationship between them (Oblak et al., 2021; Ryan et al., 2020). However, in this study, visceral fat area, rather than BMI, was most associated with DNAmAgeAccel. The results of this study are consistent with previous studies showing that visceral fat mass or visceral fat area assessed by computed tomography (CT) or dual-energy X-ray absorptiometry (DXA) is positively related to DNAmAgeAccel (*i.e.*, GrimAgeAccel) (Arpón et al., 2019; Lu et al., 2019). The differences in results with previous studies observed in BMI could be attributed to the relatively small range of BMI (20.9–26.2) of the participants in this study. However, the findings of this study represent that visceral fat area can be an important variable for estimating biological aging even in participants with relatively homogeneous BMI.

Among the blood biochemical parameters, there was a significant positive correlation between serum TG and PhenoAgeAccel, GrimAgeAccel, and FitAgeAccel, respectively, while serum HDL-C had a negative correlation with PhenoAgeAccel and GrimAgeAccel(**Figure 4a-f**). While comparing the tertiles, GrimAgeAccel in the group with the highest glucose and TG levels were approximately 1.6 years faster than in the lowest groups (**Figure 4g** and **Table S4**). In addition, PhenoAgeAccel and GrimAgeAccel of the groups with the highest HDL-C were -3.2 and -2.0 years slower than in the middle group, respectively (**Figure 4h** and **4i** and **Table S3 and S4**). The blood energy substrates glucose and TG were already shown to positively correlate with both PhenoAgeAccel and GrimAgeAccel when these clocks were developed (Lu et al., 2019; Levine et al., 2018). It is known that these substrates also correlate well with physical fitness levels and body composition, suggesting the importance of maintaining an optimal metabolic energy state for delaying biological aging. Alternately, HDL-C also negatively correlated with both PhenoAgeAccel and GrimAgeAccel (Lu et al., 2019; Levine et al., 2018). HDL-C is widely used in clinical settings, mainly as a biomarker for estimating the cardiovascular risk. However, a recent study suggested that there is a J-shaped dose-response manner between blood HDL-C levels and cardiovascular diseases and all-cause mortality (Zhong et al., 2020). Therefore, the upper limit of the blood HDL-C level that has a favorable effect on biological aging is currently unknown.

In terms of nutrients variables, relative daily intakes corrected for 1000 kcal of CHO, Fe, Cu, vitamin C, and *β*-carotene were negatively related to PhenoAgeAccel and GrimAgeAccel, respectively (**Figure 5a-5e**). Furthermore, comparisons among the tertile groups showed that the group with the highest intake of the above nutrients and micronutrients had a slower DNAmAgeAccel of 1.5 -2.6 years compared with the group with the lowest intake (**Figure 5m-5t**, **Table S3-5**). Limited evidence suggests that nutritional biomarkers in blood, such as *β*-carotene and carotenoids, are negatively associated with PhenoAgeAccel and GrimAgeAccel (Lu et al., 2019; Levine et al., 2018). In contrast, the relationship between DNAmAgeAccel and daily intake of CHO, minerals, and antioxidants largely remains unknown. The findings of this study suggest that intake of CHO and micronutrients are associated with delayed biological age progression.

Regarding smoking status, GrimAgeAccel and FitAgeAccel were 5.6 and 3.5 years faster respectively, in the current smoker group than in the non-smoker group (**Figure 6a**, **6b,** and **Table S6**). There was also a significant difference between the GrimAgeAccel of the current and past smoker groups, with a gap of 4.5 years between them (**Figure 6a** and **Table S6**). Smoking is known to be one of the major risk factors for accelerating the DNAmAgeAccel (Freni-Sterrantino et al., 2022; Kim et al., 2022; Oblak et al., 2021). The finding of this study that history of smoking is positively associated with DNAmAgeAccel is consistent with previous studies (Carroll et al., 2017). Interestingly, in our findings, the years of smoking rather than the number of cigarettes smoked correlated with DNAmAgeAccel (GrimAgeAccel: *r* = 0.478, p < 0.001; FitAgeAccel: *r* = 0.326, *p* < 0.001), (**Figure 6e-g**). Regarding the drinking status, the GrimAgeAccel was approximately 1.7 years faster in the group that drank 5-7 times a week than the group that drank once a week or less (**Figure 6c** and **Table S7**). This result is slightly different from a previous report showing a gradual U-shaped pattern between alcohol intake and GrimAgeAccel (Kim et al., 2022), but at least these findings suggest that excessive alcohol consumption is positively associated with GrimAgeAccel. Thus, avoiding smoking, especially for many years, and avoiding excessive alcohol consumption, are related to the delay of biological aging.

The sleep quality was assessed using the PSQI score, and there were no differences in any of the DNAmAgeAccel by the presence or absence of sleep disorders (**Table S8**). Limited evidence suggests that insomnia symptoms and short, irregular sleep accelerate the DNAmAgeAccel (Carroll et al., 2017; Carskadon et al., 2019). Therefore, further research is required to understand the relationship between sleep quality and DNAmAgeAccel. However, the GrimAgeAccel was approximately 1.2 years slower in the group with a morning-type chronotype than in the intermediate and moderate evening-type group (**Figure 6d** and **Table S9**). Some studies have examined the relationship between chronotype and DNA methylation patterns (Wong et al., 2015), but to our knowledge, there is no study that examines the relationship with the DNAm aging clocks. Collectively, these findings on sleep quality and chronotype suggest that maintaining sleep quality and a morning-type chronotype may contribute, at least in part, to the delay of biological aging.

There were no differences in any of the DNAmAgeAccel in the presence of lifestyle-related diseases such as hypertension and diabetes, except for dyslipidemia (**Table S10**). DNAmPhenoAge and DNAmGrimAge were developed based on aging outcomes, such as blood biochemical parameters, cancer, heart disease, and all-cause mortality (Lu et al., 2019; Levine et al., 2018); hence it is not surprising that the DNAm aging clocks are associated with mortality and above-mentioned diseases (Oblak et al., 2021). Conversely, it was suggested that there is a marginal positive relationship between DNAmAgeAccel and lifestyle-related diseases such as dyslipidemia, hypertension, and diabetes (Oblak et al., 2021). In this study, there is no strong correlation between each DNAmAgeAccel and blood biochemical parameters (**Figure 4**) and blood pressure (data is not shown), which are the criteria that are considered for the respective diseases. These results are roughly consistent with the results of previous studies (Lu et al., 2019; Levine et al., 2018). Given these findings, the current DNAmAgeAccel may be unsuitable for predicting lifestyle-related diseases with high accuracy.

The results of multiple regression analysis indicated that objectively measured physical fitness (*i.e.*, VO_2_/kg at VT and VO_2_/kg at Peak) had a relatively low contribution to DNAmAgeAccel compared to variables such as smoking, CHO intake, and serum TG levels (**Table 2**). Alternately, in this study, the VO_2_/kg at Peak values were weakly and negatively correlated with GrimAgeAccel, and biological aging in the highest VO_2peak_ group was slower than in those with the lowest group (**Figure 2c, 2e,** and **Table S4**). Thus, it is possible that the maintenance of cardiorespiratory fitness is at least partly involved in delaying the biological aging process. At present, the number of human intervention studies targeting the DNAm aging clocks are limited (Fahy et al., 2019; Fiorito et al., 2021; Fitzgerald et al., 2021; Waziry et al., 2023). Importantly, these studies suggest that long-term pharmacological or life-style interventions are effective in not only slowing down but also reversing the DNAmAgeAccel. Although the causal relationships between each lifestyle-related variable and DNAmAgeAccel are unknown, this study suggests that even in relatively homogeneous subjects, maintaining cardiorespiratory fitness and an appropriate metabolic state, sufficient intake of CHO and micronutrients, avoiding smoking and excessive alcohol consumption, and having a morning-type chronotype are associated with delayed biological aging than with chronological aging.

Future longitudinal studies should investigate the relationships between changes in lifestyle-related variables, including VO_2peak_, with a DNAmAgeAccel and aging. It is also necessary to determine whether cardiometabolic improvements induced by aerobic exercise training interventions are effective in slowing down or reversing DNAmAgeAccel. In addition, the above studies should also consider racial and sex differences in DNAmAgeAccel (Horvath et al., 2016; Tajuddin et al., 2019).

## 4. Conclusions

In conclusion, the contribution of cardiorespiratory fitness to DNAmAgeAccel was relatively low compared to lifestyle factors such as smoking. However, this study reveals a negative relationship between cardiorespiratory fitness and DNAmAgeAccel in older men.

## 5. Methods

### 5.1. Participants

The WASEDA’S Health Study is a prospective cohort study that investigates the relationships between health outcomes and sports, exercise, physical activity, and sedentary behavior among the alumni of Waseda University and their spouses aged 40 years or older (Ito et al., 2019a, 2019b; Kawamura et al., 2021; Tanisawa et al., 2022a, 2022b). The WASEDA’S Health Study consists of four cohorts (cohorts A–D) with different measurement items, and the participants selected one of the four cohorts when registering for the study (Tanisawa et al., 2022b). The study included a total of 169 men aged 65–72 years who participated in the baseline survey of Cohort D between March 2015 and March 2020, 144 of whom were included in order of measurement date from the earliest to the latest, after excluding those whose DNA sample quality did not meet the criteria (*n* = 11). The participants were briefed on the study and signed an informed consent form prior to the baseline survey. This study was approved by the Research Ethics Committee of Waseda University (approval numbers: 2014-G002 and 2018-G001). The study was conducted in accordance with the Declaration of Helsinki (1964).

### 5.2. Self-administered questionnaires, nutrients assessment

Self-administered questionnaires were used to determine the following: age (in years), smoking habits (current, former, and non-smoking), frequency of drinking (less than once a week, 2–4 times a week, more than 5 times a week). Sleep quality was assessed using PSQI score (Backhaus et al., 2002; Doi et al., 2000), with a score of 5 points or less defined as no sleep disorder and 6 points or more as having a sleep disorder. Chronotype was assessed using the Japanese version of the MEQ (Horne & Ostberg, 1976; Ishihara et al., 1986). The MEQ consisted of 19 questions about preferred sleep time and daily performance, and the scores ranged from 16 to 86. Based on the MEQ score, participants were classified into intermediate and moderate evening type (score 31–58) or morning and definite morning type (59–86). As described previously (Ito et al., 2019a, 2019b), the brief-type self-administered dietary history questionnaire (BDHQ) was also used to investigate the intake of total energy (kcal/day), protein (g/1000 kcal/day), fat (g/1000 kcal/day), carbohydrate (g/1000 kcal/day), iron (Fe, mg/1000 kcal/day), zinc (Zn, mg/1000 kcal/day), copper (Cu, mg/1000 kcal/day), manganese (Mn, mg/1000 kcal/day), vitamin A (calculated as retinol activity equivalent, *μ*gRAE/1000 kcal/day), vitamin C (mg/1000 kcal/day), α-tocopherol (mg/1000 kcal/day), and *β*-carotene (calculated as *β*-carotene equivalent, *μ*g/1000 kcal/day). The validity of the BDHQ was evaluated in previous studies (Kobayashi et al., 2011, 2012).

### 5.3. Anthropometric measurements

Height (cm) and body weight (kg) were measured using a stadiometer (YHS-200D, YAGAMI Inc., Nagoya, Japan) and an anthropometer (MC-980A, Tanita, Tokyo, Japan), respectively. Body weight was measured with light clothing and without shoes. BMI (kg/m^2^) was calculated from height and body weight measurements. Body fat (%) was measured using bioelectrical impedance analysis (MC-980A, Tanita, Tokyo, Japan). Fat free mass (kg) was calculated from the body weight and fat. Visceral fat area (cm^2^) and subcutaneous fat area (cm^2^) were measured using magnetic resonance imaging (MRI) (Signa Premier; GE Healthcare, Waukesha, WI, USA) as described previously (Usui et al., 2020). Abdominal circumference (cm) was measured to the nearest 0.1 cm at the umbilical region with an inelastic measuring tape at the end of normal expiration. Calf circumference (cm) was measured in 0.1 cm increments in the standing position; twice on each side where the circumference was the greatest. Details of the two circumferential measurements can be found in the previous studies (Kawakami et al., 2020).

### 5.4. Blood pressure and physical fitness measurements

Resting systolic and diastolic blood pressures (SBP and DBP, mmHg) were measured at least twice in the sitting position using an automatic sphygmomanometer (HEM-7250-IT; Omron Healthcare Co., Ltd., Kyoto, Japan), and the mean value was used (Tanisawa et al., 2022b). VO_2peak_ (mL/min and mL/kg/min), grip strength (kg), and leg extension power (W) were measured as previously reported (Kawamura et al., 2021). Specifically, VO_2peak_ was measured using the breath-by-breath method with a bicycle ergometer (828E, Monarch, Stockholm, Sweden). Measurements started at 30 W after 3 minutes of rest, and the load was gradually increased by 15 W per minute until exhaustion. The endpoint of the exercise stress test was defined as the plateau of oxygen uptake, or the heart rate reached approximately 90% of the predicted maximum age-specific heart rate. The exercise test was stopped if the perceived exertion’s rating (RPE) reached 18 or higher, if the subject reported that he could not continue exercising, or if the SBP reached 250 mmHg. VO_2peak_ was defined as the highest value of average oxygen uptake during 30 seconds of exercise, and VO_2_ at VT (mL/min and mL/kg/min) was also calculated. Heart rate (bpm) and blood pressure were monitored before and during exercise for the safety of the older participants. Grip strength was measured twice, alternately on the left and right hand, and the average value of right-hand was used in this study. Leg extension power was repeatedly measured five times using a recumbent leg press (Anaero Press 3500, Combi, Tokyo, Japan) and the maximum value was adopted.

### 5.5. Blood sampling and biochemical measurements

The procedure from blood collection to blood sample storage was as previously reported (Kawamura et al., 2021). Participants were briefly instructed to fast for at least 12 hours at the night before the blood collection. Venous blood was collected from a forearm vein into blood collection tubes (TERUMO, Tokyo, Japan) with or without anticoagulant (EDTA-2Na with NaF and heparin sodium) and centrifuged at 4°C for 10 minutes at 3000 rpm using a centrifuge (Model 5911, Kubota, Tokyo, Japan). The supernatant was transferred to microtubes and both plasma and serum samples were stored in a -80°C freezer until analysis. Serum samples were analyzed for insulin (*μ*U/mL), TG (mg/dL), total cholesterol (total-C, mg/dL), HDL-C (mg/dL), and low-density lipoprotein cholesterol (LDL-C, mg/dL). Plasma samples were analyzed for fasting blood glucose (glucose, mg/dL) and hemoglobin A1c (HbA1c, %). All analyses of these blood biochemical parameters were performed by BML, Inc. (Tokyo, Japan).

### 5.6. Definition of diseases

According to the guidelines of the Japan Atherosclerosis Society (Teramoto et al., 2012), dyslipidemia is defined as having at least one of the following components: fasting LDL levels ≥140 mg/dL, fasting HDL-C levels <40 mg/dL, and fasting TG levels ≥150 mg/dL. Additionally, according to the guidelines of the Japanese Society of Hypertension’s guidelines (Umemura et al., 2019), hypertension is defined as having both or either of the following components: SBP ≥140 mmHg and DBP ≥90 mmHg. According to the guidelines of the Japan Diabetes Society (Araki et al., 2020), diabetes is defined as having both or either fasting glucose ≥126 mg/dL and HbA1c ≥6.5%. Lifestyle-related diseases is defined as having one or more of the above, dyslipidemia, hypertension, or diabetes mellitus.

### 5.7. DNA extraction and epigenome-wide DNA methylation measurement

For DNA extraction, blood was drawn into collection tubes containing anticoagulant (EDTA-2Na). DNA extraction using whole blood was performed according to the manufacturer’s instructions using QIAamp DNA Midi Kit (Qiagen, Germany). Extracted DNA was dissolved in Buffer AE (10 mM Tris-Cl, 0.5 mM EDTA, pH 9.0) Prior to DNA methylation measurement, DNA samples were adjusted to a concentration of ≥ 50 ng/μL with a purity of A260/280 ranging from > 1.7–2.1. Epigenome-wide DNA methylation was measured using the same procedure as previously reported (Jokai et al., 2022). Briefly, genomic DNA bisulfite conversion was performed by using the EZ DNA Methylation Kit (Zymo Research, Irvine, CA, USA), followed by hybridization using Infinium MethylationEPIC BeadChip Kit (Illumina Inc., San Diego, CA, USA). Sample- and probe-based quality checking were performed using R version 4.0.5 and they involved the use of minfi, Meffil, and ewastools R packages (Jokai et al., 2022). In this study, no samples were excluded as all 144 DNA samples passed the criteria set by Illumina.

### 5.8. Epigenetic biomarkers

Five DNAm aging clocks, DNAmAgeHorvath (years), DNAmAgeHannum (years), DNAmPhenoAge (years), DNAmGrimAge (years), and DNAmFitAge (years), were calculated using the obtained epigenome-wide DNA methylation data as previously reported (Hannum et al., 2013; Horvath, 2013; Lu et al., 2019; McGreevy et al., 2023; Levine et al., 2018). Based on the residuals of the regression of each DNAm aging clock on chronological age, age-adjusted estimates of DNAmAgeAccel, *i.e.*, HorvathAgeAccel (years), HannumAgeAccel (years), PhenoAgeAccel (years), GrimAgeAccel (years), and FitAgeAccel (years) were calculated.

### 5.9. Statistical analysis

As descriptive data, continuous variables are presented as mean ± standard deviation (SD) and categorical variables are presented as the number of persons and percentages, respectively (Table 1 and S1). Participants were classified into tertiles (T1, T2, and T3 group) for each variable and the DNAmAgeAccel of each group was compared. Specifically, the means were compared by 1-way-ANOVA after checking homogeneity of variance for each variable. The Tukey HSD test was used for post hoc testing. Notably, the comparison of the means of the tertile groups were not adjusted for chronological age, as the effect on the results was extremely small. An unpaired student t-test was used to compare the means of the DNAmAgeAccel between the two groups. Pearson’s correlation analysis was performed for the relationships between chronological age and each DNAm aging clock, and between each variable and DNAmAgeAccel, and correlation coefficients (*r*) were presented.

Three multiple regression models were developed to identify the determinants of each DNAm aging clock, and the standard partial regression coefficients and 95% confidence intervals for the independent variables were calculated. The dependent variables in each model were PhenoAgeAccel, GrimAgeAccel, and FitAgeAccel. The candidate independent variables in each model were body fat (%), visceral fat area (cm^2^), fat free mass (kg), calf circumference (cm), VO_2_/kg at VT (mL/kg/min), VO_2_/kg at Peak (mL/kg/min), TG (mg/dL), HDL-C (mg/dL), CHO (g/1000 kcal/day), Fe (mg/1000 kcal/day), Cu (mg/1000 kcal/day), vitamin C (mg/1000 kcal/day), *β*-carotene (*μ*g/1000 kcal/day), smoking (non-smoker or past smoker or current smoker), drinking (0–1 times a week or 2–4 times a week or 5–7 times a week), and chronotype (moderate and definite morning type or intermediate and moderate evening type). The enter method was used to develop the model, and before using this method, we calculated the variance inflation factor (VIF) for each variable to avoid multicollinearity and to ensure that the VIF was less than 10. A two-sided *p* value of less than 5% was expressed as statistically significant. All statistical analyses were performed using SPSS Statistics version 26 (IBM Corporation, Chicago, IL, USA).

## Data Availability

The measurement data used to support the findings of this study is available from the corresponding author upon reasonable request.

## Acknowledgements

We would like to thank all the staff of WASEDA’S Health Study and all the participants in the experiment. We also thank Editage (https://www.editage.jp/) for English editing.

## Author’s contributions

T.K., Z.R., and K.T. conceptualized and designed the study. S.S., K.O., M.H., I. M., and K.T. conceived and supervised the study. T.K., H.T., N.N., R.K., T.I., C.U., H-K.K., K.S., K.I., S.S., and K.T. conducted the investigation. KM.M., and S.H. calculated the DNA methylation age and age acceleration. T.K., H.A., and K.T. performed statistical analyses and created the figures. T.K., Z.R., M.J., F.T., M.M., and K.T. interpreted the data. T.K. wrote the manuscript and all authors read and agreed to the published version of the manuscript.

