## Supplemental Data for "Associations between cardiorespiratory fitness and lifestyle-related factors with DNA methylation-based aging clocks in older men: WASEDA’S Health Study"

Table S1. Characteristics of the participants in categorical variables

| | ***n*** **(%)** | |
|---|---|---|
| ***Smoking status*** | **144** | |
| Non-smoker | 44 | (30.6) |
| Past smoker | 90 | (62.5) |
| Current smoker | 10 | (6.9) |
| ***Drinking status*** | **144** | |
| 0–1 times/week | 41 | (28.5) |
| 2–4 times/week | 27 | (18.7) |
| 5–7 times/week | 76 | (52.8) |
| ***Sleep disorders*** | **143** | |
| Yes | 43 | (30.0) |
| No | 100 | (70.0) |
| ***Chronotype*** | **141** | |
| Moderate and obvious morning type | 83 | (58.9) |
| Intermediate and moderate evening type | 58 | (41.1) |
| ***Dyslipidemia*** | **144** | |
| Yes | 61 | (42.4) |
| No | 83 | (57.6) |
| ***Hypertension*** | **144** | |
| Yes | 68 | (47.2) |
| No | 76 | (52.8) |
| ***Diabetes*** | **144** | |
| Yes | 20 | (13.9) |
| No | 124 | (86.1) |
| ***Lifestyle-related diseases*** | **144** | |
| Yes | 97 | (67.4) |
| No | 47 | (32.6) |

Table S2. Mean of the tertile groups in each continuous variable

| | T1 (Lowest group) | T2 (Medium group) | T3 (Highest group) | *n* |
|---|---|---|---|---|
| ***Anthropometric variables*** | | | | |
| Height (cm) | 161.5 ± 2.9 | 168.0 ± 1.6 | 174.3 ± 2.7 | 144 |
| Body weight (kg) | 58.0 ± 2.8 | 65.0 ± 2.0 | 75.4 ± 6.0 | 144 |
| BMI (kg/m$^2$) | 20.8 ± 1.0 | 23.3 ± 0.6 | 26.2 ± 1.6 | 144 |
| Body fat (%) | 15.1 ± 3.1 | 21.7 ± 1.1 | 26.9 ± 2.6 | 144 |
| Fat free mass (kg) | 47.1 ± 2.4 | 51.4 ± 1.0 | 56.9 ± 3.0 | 144 |
| Visceral fat area (cm$^2$) | 52.0 ± 18.0 | 92.4 ± 11.0 | 146.1 ± 36.4 | 139 |
| Subcutaneous fat area (cm$^2$) | 75.1 ± 17.5 | 121.0 ± 10.2 | 175.7 ± 38.5 | 139 |
| Abdominal circumference (cm) | 76.8 ± 3.8 | 84.6 ± 1.8 | 92.9 ± 4.8 | 144 |
| Calf circumference (cm) | 34.4 ± 1.0 | 36.6 ± 0.6 | 38.9 ± 1.2 | 136 |
| ***Physical fitness variables and blood pressure*** | | | | |
| $VO_2$ at VT (mL/min) | 791.9 ± 90.2 | 990.4 ± 46.6 | 1226.0 ± 158.9 | 142 |
| $VO_2$/kg at VT (mL/kg/min) | 11.9 ± 1.1 | 15.2 ± 0.9 | 18.9 ± 2.8 | 142 |
| $VO_2$ at peak (mL/min) | 1421.8 ± 132.7 | 1722.0 ± 74.5 | 2065.6 ± 162.1 | 144 |
| $VO_2$/kg at peak (mL/kg/min) | 21.8 ± 1.9 | 26.2 ± 1.0 | 31.5 ± 3.3 | 144 |
| Grip strength (kg) | 29.2 ± 3.5 | 35.4 ± 1.3 | 41.2 ± 3.1 | 143 |
| Leg extension power (W) | 766.3 ± 114.0 | 1072.1 ± 63.9 | 1354.8 ± 143.3 | 143 |
| SBP (mmHg) | 118.3 ± 10.2 | 136.7 ± 3.3 | 161.5 ± 13.8 | 144 |
| DBP (mmHg) | 71.3 ± 6.3 | 84.4 ± 2.7 | 97.2 ± 7.4 | 144 |
| ***Blood biochemical variables*** | | | | |
| Insulin ($\mu$U/mL) | 3.0 ± 0.8 | 5.6 ± 0.8 | 11.1 ± 4.2 | 144 |
| Glucose (mg/dL) | 88.8 ± 4.5 | 101.3 ± 3.8 | 123.4 ± 18.1 | 144 |
| HbA1c (%) | 5.3 ± 0.1 | 5.6 ± 0.1 | 6.2 ± 0.4 | 144 |
| TG (mg/dL) | 59.0 ± 12.9 | 99.5 ± 12.1 | 176.6 ± 65.8 | 144 |
| Total-C (mg/dL) | 178.6 ± 12.4 | 215.0 ± 9.7 | 249.1 ± 17.4 | 144 |
| HDL-C (mg/dL) | 48.1 ± 6.1 | 61.1 ± 3.5 | 81.8 ± 12.4 | 144 |
| LDL-C (mg/dL) | 93.5 ± 14.3 | 122.3 ± 7.5 | 157.6 ± 15.0 | 144 |
| ***Nutrients variables*** | | | | |
| Energy intake (kcal/day) | 1548 ± 220 | 2038 ± 138 | 2732 ± 396 | 144 |
| Protein (g/1000 kcal/day) | 34.1 ± 3.5 | 42.0 ± 1.7 | 49.7 ± 5.3 | 144 |
| Fat (g/1000 kcal/day) | 25.2 ± 2.9 | 31.3 ± 1.4 | 39.4 ± 5.6 | 144 |
| CHO (g/1000 kcal/day) | 93.2 ± 14.4 | 120.2 ± 6.2 | 140.1 ± 8.8 | 144 |
| Fe (mg/1000 kcal/day) | 3.8 ± 0.5 | 4.8 ± 0.2 | 6.1 ± 1.0 | 144 |
| Zn (mg/1000 kcal/day) | 3.8 ± 0.4 | 4.6 ± 0.2 | 5.4 ± 0.6 | 144 |
| Cu (mg/1000 kcal/day) | 0.5 ± 0.05 | 0.6 ± 0.03 | 0.8 ± 0.11 | 144 |
| Mn (mg/1000 kcal/day) | 1.2 ± 0.2 | 1.5 ± 0.1 | 2.1 ± 0.3 | 144 |
| Vitamin A ($\mu$g RAE/1000 kcal/day) | 277 ± 63 | 464 ± 51 | 807 ± 255 | 144 |
| Vitamin C (mg/1000 kcal/day) | 46.9 ± 9.5 | 68.9 ± 6.0 | 107.0 ± 30.7 | 144 |
| $\alpha$-tocopherol (mg/1000 kcal/day) | 3.5 ± 0.4 | 4.5 ± 0.3 | 5.7 ± 1.0 | 144 |
| $\beta$-carotene ($\mu$g/1000 kcal/day) | 1221 ± 325 | 2088 ± 220 | 3695 ± 1570 | 144 |

Data are mean ± standard deviation (SD). BMI, body mass index; VT, ventilatory threshold; SBP, systolic blood pressure; DBP, diastolic blood pressure; HbA1c, Hemoglobin A1c; TG, triglyceride; Total-C, total cholesterol; HDL-C, high-density lipoprotein cholesterol; LDL-C, low-density lipoprotein cholesterol; CHO, carbohydrate; Fe, iron; Zn, zinc; Cu, copper; Mn, manganese; Accel, acceleration.

Table S3. PhenoAgeAccelration of the tertile groups in each continuous variable

| | T1 (Lowest group) | T2 (Medium group) | T3 (Highest group) | *p* value |
|---|---|---|---|---|
| ***Anthropometric variables*** | | | | |
| Height (cm) | 0.05 ± 5.54 | 1.03 ± 5.39 | -1.08 ± 5.38 | 0.167 |
| Body weight (kg) | -0.81 ± 5.42 | 1.07 ± 5.08 | -0.26 ± 5.82 | 0.225 |
| BMI (kg/m$^2$) | -0.87 ± 5.49 | 0.61 ± 5.11 | 0.26 ± 5.78 | 0.384 |
| Body fat (%) | -1.80 ± 4.65 | 0.77 ± 5.55 | 1.03 ± 5.80 * | **0.019** |
| Fat free mass (kg) | 0.43 ± 5.34 | -0.05 ± 5.68 | -0.38 ± 5.46 | 0.771 |
| Visceral fat area (cm$^2$) | -0.79 ± 5.40 | -0.96 ± 4.71 | 1.85 ± 5.96 † | **0.022** |
| Subcutaneous fat area (cm$^2$) | -0.23 ± 6.32 | -0.24 ± 4.31 | 0.56 ± 5.73 | 0.731 |
| Abdominal circumference (cm) | -0.88 ± 5.38 | 0.38 ± 6.02 | 0.50 ± 4.97 | 0.397 |
| Calf circumference (cm) | 0.25 ± 5.99 | -0.60 ± 4.87 | 0.29 ± 5.86 | 0.694 |
| ***Physical fitness variables*** | | | | |
| $VO_2$ at VT (mL/min) | 0.63 ± 5.32 | -1.13 ± 5.81 | 0.05 ± 4.66 | 0.258 |
| $VO_2$/kg at VT (mL/kg/min) | 0.53 ± 5.94 | 0.71 ± 5.50 | -1.12 ± 4.91 | 0.203 |
| $VO_2$ at peak (mL/min) | 0.38 ± 5.38 | -0.63 ± 6.29 | 0.24 ± 4.67 | 0.623 |
| $VO_2$/kg at peak (mL/kg/min) | 0.53 ± 5.94 | 0.64 ± 5.46 | -1.18 ± 4.88 | 0.189 |
| Grip strength (kg) | 0.91 ± 5.32 | -0.35 ± 4.83 | -0.56 ± 6.17 | 0.361 |
| Leg extension power (W) | 0.25 ± 4.87 | 1.42 ± 6.12 | -1.67 ± 5.05 † | **0.021** |
| ***Blood biochemical variables*** | | | | |
| Insulin ($\mu$U/mL) | -0.75 ± 4.35 | 0.31 ± 6.70 | 0.44 ± 5.13 | 0.509 |
| Glucose (mg/dL) | -0.25 ± 4.82 | 0.24 ± 4.66 | 0.01 ± 6.77 | 0.912 |
| HbA1c (%) | -0.27 ± 4.85 | 0.54 ± 5.55 | -0.27 ± 6.01 | 0.703 |
| TG (mg/dL) | -0.17 ± 5.17 | -0.95 ± 5.78 | 1.12 ± 5.35 | 0.173 |
| Total-C (mg/dL) | 0.50 ± 4.65 | 0.33 ± 6.09 | -0.83 ± 5.58 | 0.434 |
| HDL-C (mg/dL) | 0.06 ± 5.52 | 1.65 ± 5.37 | -1.71 ± 5.10 †† | **0.010** |
| LDL-C (mg/dL) | 0.73 ± 5.05 | -0.91 ± 5.49 | 0.18 ± 5.82 | 0.326 |
| ***Nutrients variables*** | | | | |
| Energy intake (kcal/day) | -0.01 ± 5.89 | 0.59 ± 5.69 | -0.57 ± 4.82 | 0.586 |
| Protein (g/1000kcal/day) | 0.05 ± 5.55 | 0.41 ± 5.58 | -0.46 ± 5.36 | 0.739 |
| Fat (g/1000kcal/day) | -0.19 ± 5.83 | 0.27 ± 5.56 | -0.08 ± 5.09 | 0.913 |
| CHO (g/1000kcal/day) | -0.35 ± 5.15 | 0.85 ± 5.91 | -0.50 ± 5.33 | 0.419 |
| Fe (mg/1000kcal/day) | 1.64 ± 5.97 | -0.63 ± 5.09 | -1.01 ± 5.02 * | **0.036** |
| Zn (mg/1000kcal/day) | 0.78 ± 5.42 | -0.35 ± 5.22 | -0.43 ± 5.79 | 0.480 |
| Cu (mg/1000kcal/day) | 1.29 ± 5.90 | 0.05 ± 5.36 | -1.34 ± 4.89 | 0.061 |
| Mn (mg/1000kcal/day) | 0.12 ± 5.78 | -0.45 ± 5.96 | 0.32 ± 4.66 | 0.777 |
| Vitamin A ($\mu$g RAE/1000kcal/day) | 0.003 ± 6.65 | 0.70 ± 3.93 | -0.71 ± 5.50 | 0.448 |
| Vitamin C (mg/1000kcal/day) | 0.76 ± 6.17 | 0.43 ± 5.66 | -1.19 ± 4.33 | 0.175 |
| $\alpha$-tocopherol (mg/1000kcal/day) | 0.31 ± 6.32 | 0.52 ± 5.56 | -0.83 ± 4.37 | 0.433 |
| $\beta$-carotene ($\mu$g/1000kcal/day) | 1.32 ± 6.42 | -0.33 ± 4.90 | -0.99 ± 4.77 | 0.102 |

Data are mean ± standard deviation (SD). Significantly different from T1 at $p < 0.05$ is indicated by *. Significantly different from T2 at $p < 0.05$ and 0.01 are indicated by †, ††, respectively. BMI, body mass index; VT, ventilatory threshold; TG, triglyceride; Total-C, total cholesterol; HDL-C, high-density lipoprotein cholesterol; LDL-C, low-density lipoprotein cholesterol; MCV, mean corpuscular volume; MCH, $\gamma$-GTP, gamma-glutamyltransferase; ApoA-1, Apolipoprotein A-1; ApoB, Apolipoprotein B; CHO, carbohydrate; Fe, iron; Zn, zinc; Cu, copper; Mn, manganese.

Table S4. GrimAgeAcceleration of the tertile groups in each continuous variable.

| | **T1 (Lowest group)** | **T2 (Medium group)** | **T3 (Highest group)** | *p* value |
|---|---|---|---|---|
| ***Anthropometric variables*** | | | | |
| Height (cm) | 0.06 ± 2.93 | 0.14 ± 3.71 | -0.20 ± 2.81 | 0.857 |
| Body weight (kg) | -0.25 ± 2.84 | 0.37 ± 3.04 | -0.12 ± 3.58 | 0.599 |
| BMI (kg/m$^2$) | -0.87 ± 2.62 | 0.97 ± 2.94 * | -0.09 ± 3.62 | **0.015** |
| Body fat (%) | -0.79 ± 2.59 | 0.28 ± 3.46 | 0.51 ± 3.27 | 0.101 |
| Fat free mass (kg) | 0.17 ± 2.93 | -0.11 ± 2.98 | -0.05 ± 3.58 | 0.899 |
| Visceral fat area (cm$^2$) | -0.78 ± 3.00 | -0.11 ± 2.66 | 0.99 ± 3.56 * | **0.023** |
| Subcutaneous fat area (cm$^2$) | -0.01 ± 2.83 | -0.29 ± 3.30 | 0.37 ± 3.38 | 0.611 |
| Abdominal circumference (cm) | -0.31 ± 2.89 | 0.08 ± 3.45 | 0.23 ± 3.15 | 0.690 |
| Calf circumference (cm) | 0.43 ± 2.77 | -0.23 ± 3.52 | -0.37 ± 3.26 | 0.441 |
| ***Physical fitness variables*** | | | | |
| $VO_2$ at VT (mL/min) | 0.83 ± 3.06 | -0.59 ± 3.25 | -0.05 ± 3.10 | 0.054 |
| $VO_2$/kg at VT (mL/kg/min) | 0.94 ± 3.48 | -0.24 ± 3.09 | -0.62 ± 2.73 * | **0.042** |
| $VO_2$ at peak (mL/min) | 0.49 ± 3.07 | -0.35 ± 3.29 | -0.14 ± 3.11 | 0.406 |
| $VO_2$/kg at peak (mL/kg/min) | 0.94 ± 3.48 | -0.28 ± 3.07 | -0.65 ± 2.70 * | **0.035** |
| Grip strength (kg) | 0.87 ± 3.24 | -0.26 ± 2.86 | -0.62 ± 3.23 | 0.053 |
| Leg extension power (W) | -0.03 ± 2.60 | 0.45 ± 3.61 | -0.36 ± 3.23 | 0.468 |
| ***Blood biochemical variables*** | | | | |
| Insulin ($\mu$U/mL) | -0.69 ± 2.61 | 0.20 ± 3.47 | 0.49 ± 3.27 | 0.160 |
| Glucose (mg/dL) | -0.83 ± 3.07 | 0.04 ± 2.81 | 0.79 ± 3.42 * | **0.041** |
| HbA1c (%) | -0.30 ± 3.50 | -0.33 ± 2.52 | 0.62 ± 3.33 | 0.241 |
| TG (mg/dL) | -0.64 ± 2.39 | -0.32 ± 3.33 | 0.96 ± 3.47 * | **0.030** |
| Total-C (mg/dL) | -0.41 ± 3.18 | 0.04 ± 3.29 | 0.37 ± 3.01 | 0.484 |
| HDL-C (mg/dL) | 0.11 ± 3.62 | 0.96 ± 3.06 | -1.07 ± 2.39 †† | **0.006** |
| LDL-C (mg/dL) | -0.10 ± 3.20 | -0.51 ± 3.32 | 0.61 ± 2.89 | 0.218 |
| ***Nutrients variables*** | | | | |
| Energy intake (kcal/day) | -0.26 ± 3.44 | 0.44 ± 3.14 | -0.18 ± 2.88 | 0.502 |
| Protein (g/1000kcal/day) | 0.11 ± 3.34 | -0.20 ± 3.07 | 0.09 ± 3.12 | 0.868 |
| Fat (g/1000kcal/day) | -0.09 ± 3.09 | 0.16 ± 3.13 | -0.07 ± 3.31 | 0.916 |
| CHO (g/1000kcal/day) | 0.58 ± 3.54 | 0.36 ± 3.08 | -0.94 ± 2.64 * | **0.039** |
| Fe (mg/1000kcal/day) | 1.22 ± 3.51 | -0.73 ± 2.79 ** | -0.49 ± 2.83 * | **0.004** |
| Zn (mg/1000kcal/day) | 0.77 ± 3.24 | -0.45 ± 3.12 | -0.32 ± 3.03 | 0.117 |
| Cu (mg/1000kcal/day) | 0.91 ± 3.48 | 0.22 ± 3.10 | -1.14 ± 2.53 ** | **0.005** |
| Mn (mg/1000kcal/day) | 0.60 ± 3.46 | -0.68 ± 2.90 | 0.08 ± 3.02 | 0.135 |
| Vitamin A ($\mu$g RAE/1000kcal/day) | 0.12 ± 3.28 | 0.61 ± 3.15 | -0.73 ± 2.95 | 0.110 |
| Vitamin C (mg/1000kcal/day) | 0.76 ± 3.53 | 0.06 ± 3.21 | -0.81 ± 2.52 * | **0.050** |
| $\alpha$-tocopherol (mg/1000kcal/day) | 0.83 ± 3.39 | -0.11 ± 2.86 | -0.72 ± 3.07 | 0.053 |
| $\beta$-carotene ($\mu$g/1000kcal/day) | 1.13 ± 3.54 | 0.10 ± 2.63 | -1.23 ± 2.83 *** | **0.0009** |

Data are mean ± standard deviation (SD). Significantly different from T1 at $p < 0.05$, $p < 0.01$ and $p < 0.001$ are indicated by *, **, ***, respectively. Significantly different from T2 at $p < 0.01$is indicated by ††. BMI, body mass index; VT, ventilatory threshold; TG, triglyceride; Total-C, total cholesterol; HDL-C, high-density lipoprotein cholesterol; LDL-C, low-density lipoprotein cholesterol; MCV, mean corpuscular volume; MCH, mean corpuscular hemoglobin; $\gamma$-GTP, gamma-glutamyltransferase; ApoA-1, Apolipoprotein A-1; ApoB, Apolipoprotein B; CHO, carbohydrate; Fe, iron; Zn, zinc; Cu, copper; Mn, manganese.

Table S5. DNAmFitAgeAcceleration of the tertile groups in each continuous variable

| | T1 (Lowest group) | T2 (Medium group) | T3 (Highest group) | *p* value |
|---|---|---|---|---|
| ***Anthropometric variables*** | | | | |
| Height (cm) | -0.14 ± 3.03 | 0.89 ± 3.86 | -1.11 ± 2.60 †† | **0.011** |
| Body weight (kg) | 0.001 ± 3.15 | 0.53 ± 3.34 | -0.89 ± 3.28 | 0.102 |
| BMI (kg/m$^2$) | -0.15 ± 3.26 | 0.39 ± 3.20 | -0.59 ± 3.39 | 0.346 |
| Body fat (%) | -0.46 ± 3.17 | 0.02 ± 3.51 | 0.09 ± 3.21 | 0.680 |
| Fat free mass (kg) | 0.56 ± 3.20 | -0.09 ± 3.40 | -0.82 ± 3.17 | 0.120 |
| Visceral fat area (cm$^2$) | -0.19 ± 3.49 | -0.70 ± 2.81 | 0.64 ± 3.49 | 0.144 |
| Subcutaneous fat area (cm$^2$) | 0.55 ± 3.52 | -0.57 ± 2.92 | -0.25 ± 3.40 | 0.248 |
| Abdominal circumference (cm) | 0.23 ± 3.47 | -0.34 ± 3.38 | -0.24 ± 3.03 | 0.667 |
| Calf circumference (cm) | 0.82 ± 2.98 | -0.45 ± 3.13 | -0.99 ± 3.57 * | **0.026** |
| ***Physical fitness variables*** | | | | |
| $VO_2$ at VT (mL/min) | 0.55 ± 3.10 | -0.76 ± 3.54 | -0.29 ± 3.06 | 0.137 |
| $VO_2$/kg at VT (mL/kg/min) | -0.38 ± 3.55 | -0.21 ± 3.61 | -0.46 ± 2.65 | 0.450 |
| $VO_2$ at peak (mL/min) | 0.40 ± 3.21 | -0.46 ± 3.70 | -0.30 ± 2.89 | 0.405 |
| $VO_2$/kg at peak (mL/kg/min) | 0.38 ± 3.55 | -0.22 ± 3.57 | -0.51 ± 2.66 | 0.405 |
| Grip strength (kg) | 0.52 ± 3.47 | -0.31 ± 3.22 | -0.57 ± 3.12 | 0.237 |
| Leg extension power (W) | -0.09 ± 2.90 | 0.56 ± 3.92 | -0.85 ± 2.91 | 0.115 |
| ***Blood biochemical variables*** | | | | |
| Insulin ($\mu$U/mL) | -0.65 ± 2.62 | 0.34 ± 4.18 | -0.04 ± 2.83 | 0.331 |
| Glucose (mg/dL) | -0.58 ± 3.25 | -0.31 ± 3.08 | 0.53 ± 3.49 | 0.225 |
| HbA1c (%) | -0.40 ± 3.52 | -0.16 ± 3.04 | 0.20 ± 3.31 | 0.664 |
| TG (mg/dL) | -0.79 ± 2.87 | -0.36 ± 3.48 | 0.79 ± 3.34 | 0.050 |
| Total-C (mg/dL) | -0.74 ± 3.15 | -0.39 ± 3.28 | 0.77 ± 3.30 | 0.060 |
| HDL-C (mg/dL) | -0.20 ± 3.84 | 0.56 ± 3.06 | -0.72 ± 2.82 | 0.157 |
| LDL-C (mg/dL) | -0.42 ± 3.12 | -0.57 ± 3.35 | 0.64 ± 3.31 | 0.144 |
| ***Nutrients variables*** | | | | |
| Energy intake (kcal/day) | -0.38 ± 3.59 | 0.28 ± 3.21 | -0.25 ± 3.07 | 0.586 |
| Protein (g/1000kcal/day) | 0.25 ± 3.35 | -0.45 ± 3.50 | -0.16 ± 3.03 | 0.580 |
| Fat (g/1000kcal/day) | -0.30 ± 3.24 | -0.23 ± 3.30 | 0.17 ± 3.36 | 0.753 |
| CHO (g/1000kcal/day) | 0.05 ± 3.12 | 0.57 ± 3.49 | -0.98 ± 3.12 | 0.062 |
| Fe (mg/1000kcal/day) | 1.03 ± 3.06 | -0.76 ± 3.35 * | -0.62 ± 3.20 * | **0.011** |
| Zn (mg/1000kcal/day) | 0.58 ± 3.12 | -0.50 ± 3.26 | -0.43 ± 3.43 | 0.200 |
| Cu (mg/1000kcal/day) | 0.97 ± 3.11 | 0.05 ± 3.21 | -1.37 ± 3.17 ** | **0.002** |
| Mn (mg/1000kcal/day) | 0.32 ± 3.36 | -0.62 ± 2.92 | -0.05 ± 3.54 | 0.374 |
| Vitamin A ($\mu$g RAE/1000kcal/day) | 0.08 ± 3.46 | 0.15 ± 2.72 | -0.59 ± 3.63 | 0.479 |
| Vitamin C (mg/1000kcal/day) | 0.02 ± 3.44 | 0.33 ± 3.50 | -0.70 ± 2.87 | 0.292 |
| $\alpha$-tocopherol (mg/1000kcal/day) | 0.38 ± 3.29 | -0.32 ± 3.64 | -0.43 ± 2.90 | 0.426 |
| $\beta$-carotene ($\mu$g/1000kcal/day) | 0.69 ± 3.47 | -0.12 ± 2.91 | -0.93 ± 3.32 | 0.054 |

Data are mean ± standard deviation (SD). Significantly different from T1 at $p < 0.05$, $p < 0.01$ are indicated by *, **, respectively. Significantly different from T2 at $p < 0.01$ is indicated by ††. BMI, body mass index; VT, ventilatory threshold; TG, triglyceride; Total-C, total cholesterol; HDL-C, high-density; lipoprotein cholesterol; LDL-C, low-density lipoprotein cholesterol; MCV, mean corpuscular volume; MCH, mean corpuscular hemoglobin; $\gamma$-GTP, gamma-glutamyltransferase; ApoA-1, Apolipoprotein A-1; ApoB, Apolipoprotein B; CHO, carbohydrate; Fe, iron; Zn, zinc; Cu, copper; Mn, manganese.

Table S6. DNA methylation age acceleration of the smoking status

| | **T1** (n = 44) | **T2** (n = 90) | **T3** (n = 10) | *p* value |
|---|---|---|---|---|
| PhenoAgeAccel | 0.18 ± 5.85 | -0.20 ± 5.21 | 1.06 ± 6.49 | 0.764 |
| GrimAgeAccel | -1.24 ± 2.40 | 0.12 ± 3.03 *** | 4.35 ± 3.38 *** † | **< 0.001** |
| FitAgeAccel | -0.95 ± 3.34 | -0.002 ± 3.19 | 2.50 ± 2.53 ** | **0.009** |

Data are mean ± standard deviation (SD). Significantly different from T1 at $p < 0.01$, $p < 0.001$, are indicated by **, *** respectively. Significantly different from T2 at p < 0.05 is indicated by †. T1: non-smokers, T2: past smokers, T3: current smokers. Accel, acceleration.

Table S7. DNA methylation age acceleration of the drinking status

| | **T1** (n = 41) | **T2** (n = 27) | **T3** (n = 76) | *p* value |
|---|---|---|---|---|
| PhenoAgeAccel | -0.38 ± 3.85 | -0.36 ± 6.49 | 0.33 ± 5.86 | 0.744 |
| GrimAgeAccel | -0.97 ± 2.66 | -0.52 ± 3.36 | 0.71 ± 3.19* | **0.013** |
| FitAgeAccel | -1.00 ± 2.83 | 0.79 ± 4.02 | 0.28 ± 3.18 | 0.123 |

Data are mean ± standard deviation (SD). Significantly different from T1 at $p < 0.05$ is indicated by *. T1: 0-1 time a week; T2: 2-4 times a week; T3: 5-7 times a week. Accel, acceleration.

Table S8. DNA methylation age acceleration of the Pittsburgh Sleep Quality Index score

| | Sleep disorders | | *p* value |
|---|---|---|---|
| | Yes (*n* =43) | No (*n* =100) | |
| PhenoAgeAccel | 0.24 ± 5.68 | -0.10 ± 5.43 | 0.736 |
| GrimAgeAccel | -0.36 ± 3.42 | 0.02 ± 3.07 | 0.921 |
| FitAgeAccel | -0.19 ± 3.60 | -0.08 ± 3.17 | 0.859 |

Data are mean ± standard deviation (SD). Accel, acceleration. Sleep disorder yes: 6 points or more; sleep disorder no: 5 points or less.

Table S9. DNA methylation age acceleration of the Morningness/Eveningness Questionnaire score

| | Moderate and definite morning type (*n* =58) | Intermediate and moderate evening type (*n* = 83) | *p* value |
|---|---|---|---|
| PhenoAgeAccel | -0.52 ± 5.33 | 0.46 ± 5.32 | 0.287 |
| GrimAgeAccel | -0.51 ± 3.06 | 0.65 ± 3.23 | **0.033** |
| FitAgeAccel | -0.59 ± 3.04 | 0.42 ± 3.51 | 0.072 |

Data are mean ± standard deviation (SD). Accel, acceleration. Moderate and obvious morning type: 59-86 points; Intermediate and moderate nocturnal type: 31-58 points.

Table S10. DNA methylation age acceleration of each disease

| | Yes | No | *p* value |
|---|---|---|---|
| ***Dyslipidemia*** | *n = 61* | *n = 83* | |
| PhenoAgeAccel | 0.79 ± 5.64 | -0.58 ± 5.30 | 0.135 |
| GrimAgeAccel | 1.08 ± 3.36 | -0.79 ± 2.76 | **0.0004** |
| FitAgeAccel | 0.94 ± 3.41 | -0.90 ± 2.98 | **0.0007** |
| ***Hypertension*** | *n* = 68 | *n* = 76 | |
| PhenoAgeAccel | -0.20 ± 5.69 | 0.18 ± 5.29 | 0.885 |
| GrimAgeAccel | 0.40 ± 3.35 | -0.36 ± 2.95 | 0.159 |
| FitAgeAccel | 0.13 ± 3.49 | -0.35 ± 3.09 | 0.227 |
| ***Diabetes*** | *n* = 20 | *n* = 124 | |
| PhenoAgeAccel | 1.83 ± 6.22 | -0.30 ± 5.30 | 0.106 |
| GrimAgeAccel | 0.73 ± 3.30 | -0.12 ± 3.13 | 0.264 |
| FitAgeAccel | 1.10 ± 3.07 | -0.32 ± 3.29 | 0.072 |
| ***Lifestyle-related diseases*** | *n* = 97 | *n* = 47 | |
| PhenoAgeAccel | 0.25 ± 5.48 | -0.51 ± 5.47 | 0.441 |
| GrimAgeAccel | 0.26 ± 3.21 | -0.54 ± 3.00 | 0.156 |
| FitAgeAccel | 0.18 ± 3.34 | -0.75 ± 3.10 | 0.112 |

Data are mean ± standard deviation (SD). Accel, acceleration.
